# Erythropoietin single nucleotide polymorphisms are associated with anemia and dyslipidemia in a cardiovascular disease population

**DOI:** 10.1101/2024.09.24.24314330

**Authors:** Victoria Northrup, Ashley L. Eadie, Victoria L. Nelson, Kenneth D’Souza, Ansar Hassan, Thomas Pulinilkunnil, Petra Kienesberger, Jean-Francois Légaré, Jeremy A. Simpson, Keith R. Brunt

**Author notes:** Corresponding Author: Dr. Keith R. Brunt, 100 Tucker Park Road, Box 5050, Saint John, NB, Canada, E2L 4L5, 1-(, @prof_brunt (social media).

## Abstract

Human erythropoietin (EPO) is an essential erythropoietic cytokine and recombinant biological therapeutic in anemia in chronic kidney disease. Single nucleotide polymorphisms (SNPs) in EPO have been linked to anemia and diabetic microvascular complications but no other cardiovascular comorbidities like dyslipidemia or hypertension. Here identified a clinical cohort of cardiovascular patients with anemia, dyslipidemia, hypertension, type-2 diabetes, or heart failure with preserved ejection fraction. Sanger sequencing was used for genotyping three EPO SNPs (rs1617640, rs507392 and rs551238), which were compared to clinical outcomes with SNPstat and clinical parameters profiles by student t-test. Nonstandard clinical chemistry measures of plasma EPO, hepcidin, transferrin, and ferritin were determined by ELISA. In the additive model, the C allele of rs1617640 was associated with lower EPO and anemia. In the recessive model, the SNP rs507392 genotype of GG was associated with dyslipidemia, which was associated with lower EPO in plasma and elevated total cholesterol and low-density lipoproteins. Interestingly, the G allele of rs507392 was found to be associated with hypertension, but only in females. Here we show that the SNPs in EPO are a potential risk factor for anemia, dyslipidemia, and hypertension in cardiovascular disease patients.

## Introduction

Cardiovascular disease is one of the leading causes of death worldwide[1], with multiple contributing risk factors like anemia, including some linked to erythropoietin (EPO) or the use of erythropoietic stimulating agents.[2] Heart failure with preserved ejection fraction (HFpEF), is an especially challenging multimorbid condition featuring anemia, hypertension, type-2 diabetes mellitus (T2DM), obesity, or chronic kidney disease. The underlying cause of HFpEF and optimal medical management remains a challenge[1]. Microvascular complications are typically reported in HFpEF[3], and anemia in heart failure is generally associated with higher mortality,[4] severity, and hospital admission rates[5,6]. HFpEF is also more common in females, particularly those who are older, obese, and with a higher New York Heart Association (NYHA) classification. Microvascular complications in diabetes and anemia have been linked to EPO but have not previously been investigated in HFpEF or its multimorbid feature comorbidities.

EPO is a master regulator of hematopoiesis and erythropoiesis, with secondary roles in cytoprotection and cellular metabolism.[7] Research on EPO has been extensive, yet clinical variation in EPO expression in health and disease and the underlying secondary mechanisms are poorly understood. A significant source of clinical variation in hormones like EPO can occur with haplotypes or single nucleotide polymorphisms (SNP). The entire gene of EPO has been known for decades, yet just three SNP have been identified as clinically relevant rs1617640(promoter), rs507392(intron) and rs551238(downstream). Genome-wide association studies (GWAS) and expression quantitative trait loci (eQTL) analysis have previously established that promoter SNP (rs1617640) is associated with an alteration in serum EPO,[8] a finding confirmed in healthy blood donors[9]. The promoter SNP (rs1617640) has also been linked to diabetic microvascular complications[10–18], anemia in T2DM[19], and renal dysfunction after cardiac surgery[20] (Supplemental Table 1). In intron 3, SNP rs507392 was linked to diabetic microvascular complications [10,12,13,21] and overall higher mortality in diabetics [9]. SNP rs551238 (downstream of *EPO*) has also been associated with microvascular complications[9,10,12–15,21,22] and higher hematocrit in healthy blood donors [9] (Supplemental Table 1). Together, this suggests a potential link between microvascular complications, hematocrit, and *EPO* SNPs. These *EPO* SNPs have also been associated with EPO serum levels[18]. In CKD, a decrease in EPO is associated with anemia[23], whereas in other forms of anemia, EPO can be increased up to 100-fold, suggesting insensitivity in molecular signaling.[24] Secondary risks to CVD might also be linked to EPO independent of anemia, especially classic risk factors of CVD. For example, dyslipidemia is a salient risk factor for CVD characterized by abnormal serum cholesterol levels, triglycerides and related lipoproteins[25]. In mice treated with recombinant EPO (rhEPO), triglyceride levels were decreased[26] and lipoprotein lipase (in lipogenesis) was elevated.[26,27] In humans, treatment with rhEPO is associated with lower triglyceride levels and cholesterol in end-stage renal failure.[28]

Although anemia, dyslipidemia, T2DM, hypertension are risk factors for CVD and multimorbid features of HFpEF with evidence of being modifiable by rhEPO—only anemia has been linked to *EPO* SNPs to date.[19] Here, we hypothesize that three *EPO* SNPs— rs1617640(promoter), rs507292(intron) and rs551238(downstream)—correlate with EPO plasma concentrations and are associated with either anemia, dyslipidemia, T2DM, hypertension and/or HFpEF in a clinical population requiring surgical interventions for CVD.

## Methods

### Genomic DNA extraction

Genomic DNA (gDNA) was isolated from the buffy coat of blood collected during the impact of Obesity on Postoperative Outcomes following cardiac Surgery (OPOS) study (http://clinicaltrials.gov/study/NCT0324891). The OPOS cohort consisted of participants aged 18-75 years old who underwent elective cardiac surgery at either the New Brunswick Heart Center in New Brunswick, Canada or the Maritime Heart Centre in Nova Scotia, Canada.[29] The OPOS study and related biomarker discovery was approved by the Horizon Health Network Research Ethics Board (#RS20142006: RS100835). The buffy coat was separated from blood, and gDNA was isolated using either DNAzol^®^ reagent (GibcoBRL cat # 10503-027) or QIAamp^®^ DNA blood mini (Qiagen cat # 51104) kits (following the manufacturer’s protocol). DNA was quantified by spectroscopy on a BioTek Synergy ™ H4 Hybrid Microplate reader using the 260:280 nm ratio by the Gen5 software (BioTek), and the average concentration of technical duplicates was used.

### Primer design

Primers were designed using Oligo 7 software (Table 1).[30] Primers designed for gDNA sequencing included M13 sequences[31] to facilitate Sanger sequencing and M13 primers were used in sequencing reactions.

**Table 1.**
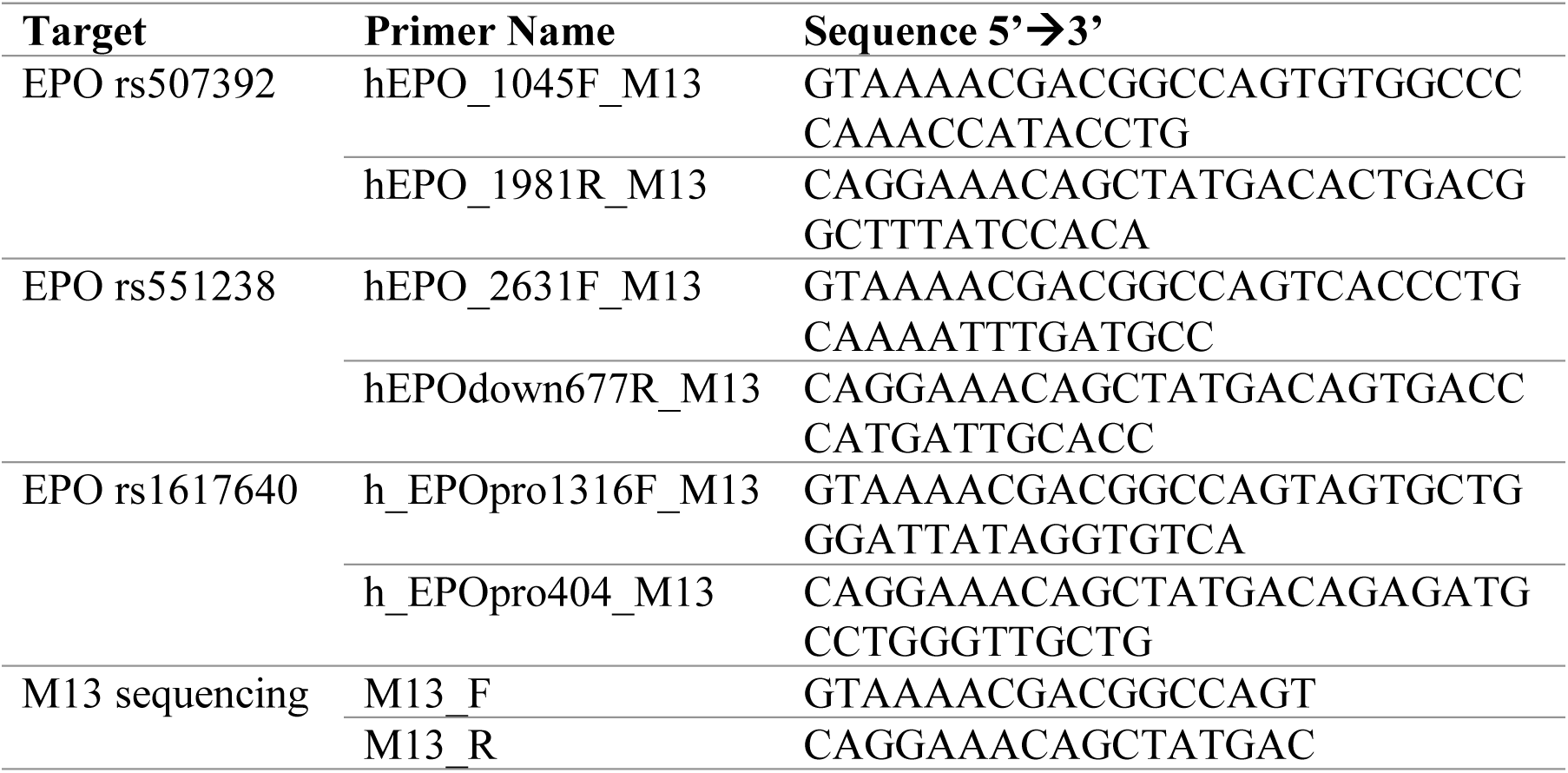
Primers.

### Polymerase Chain Reaction

End-point polymerase chain reaction (PCR) was performed on gDNA using AmpliTaq Gold^™^ 360 Master Mix (Applied Biosystems cat # 4398881). PCR reactions consisted of 1 X AmpliTaq Gold™ 360 Master Mix, 0.3 µM of forward and reverse primers, 4 µg of Bovine Serum Albumin (BSA, Thermo Scientific), 50 ng of gDNA and nuclease-free water up to 15 µL. Reactions were amplified on a Mastercycler Nexus Gradient Thermocycler (Eppendorf) using the cycling conditions listed in Table 2.

**Table 2.** Cycling conditions for gDNA end-point PCR.

| Step | h_EPOpro1316F_M13,<br>h_EPOpro404R_M13 | hEPO_1045F_M13,<br>hEPO_1981R_M13 | hEPO_2631F_M13,<br>hEPOdown677R_M13 |
| --- | --- | --- | --- |
| **Initial Denaturation** | 10 min @ 95°C | 10 min @ 95°C | 10 min @ 95°C |
| **Denaturation** | 30 sec @ 95°C | 30 sec @ 95°C | 30 sec @ 95°C |
| **Annealing** | 30 sec @ 58°C | 30 sec @ 58°C | 30 sec @ 58°C |
| **Extension** | 45 sec @ 72°C | 60 sec @ 72°C | 90 sec @ 72°C |
| **Cycles** | 35 | 35 | 35 |
| **Final Extension** | 10 min @ 72°C | 10 min @ 72°C | 10 min @ 72°C |

Min=minutes, sec=seconds @=at

PCR products were verified using 7500 Agilent DNA Chips (Agilent cat # 5067-1506) on the Bioanalyzer 2100 system (Agilent). PCR products were cleaned up using 2 µL of ExoSAP-IT™ (Applied Biosystems cat # 78200) in 5 μL of PCR product and incubating at 37°C for 15 minutes. ExoSAP-IT™ was inactivated at 80°C for 15 minutes.

### Sanger Sequencing

Sequencing reactions were prepared using a BigDye™ Terminator v3.1 Cycle Sequencing Kit (Applied Biosystems cat# 4337457). The cleaned PCR product was added to a reaction of 1X Sequencing buffer, 1X BigDye™ MasterMix, and 2μM primer of M13 sequencing primers.[31] Sequencing reactions were incubated at 96°C for 1 minute and 25 cycles of 96°C for 10 seconds, 50°C for 5 seconds and 60°C for 4 minutes.

BigDye™ Xterminator beads (Applied Biosystems cat # 4376486) were added to the sequencing reaction and vortexed at 2000 rpm for 25 minutes. Samples were centrifuged for 2 minutes at 2000 X rcf and let rest for 2 minutes before centrifuging again at 2000 X rcf for 30 seconds, followed by another 30 seconds of rest to ensure that beads were sedimented. Then, 20μL of the sequencing reaction (without beads) was transferred to a 96-well plate and loaded onto the 3500 Genetic Analyzer (Applied Biosystems) for sequencing.

### Sanger Sequencing Analysis

Sequences were analyzed using BioEdit Sequences Alignment Editor software.[32] The forward and reverse sequences were aligned using the ClustalW Multiple alignment tool to generate the consensus sequence for each product. Sequences were submitted to NCBI Blast to validate correct target amplification and were compared to the NCBI reference sequence NG_021471.2 to identify variants. A variant was defined as any base calling that differed from the reference sequence.

### SNP analysis

SNPStats (www.snpstats.net)[33] was used to determine allele and genotype frequencies, test for Hardy-Weinberg equilibrium, assess SNP response using logistic regression in multiple inheritance models (co-dominant, dominant, recessive, over-dominant and log-additive), linkage disequilibrium statistics, and for haplotype analysis. Analysis used sex, age, and body mass index (BMI) as co-variants. A p-value < 0.05 was deemed significant.

Clinical findings were obtained from the OPOS BioBank (storing retrievable databases of de-identified data abstracted from patient records). Comparisons of clinical values with genotype were performed using a student t-test to determine the p-value (<0.05 significant). A Kolmogorov-Smirnov test in GraphPad was used to confirm normality. A ROUT (Q=1) test in GraphPad was used to determine outliers; outliers were still presented and identified by encirclement in figures for clinical relevance but were otherwise excluded from group testing.

### Enzyme-linked immunosorbent assays

Plasma samples collected from OPOS [29] were used for enzyme-linked immunosorbent assays (ELISA) for EPO (Human EPO ELISA kit, Invitrogen Cat # BMS2035-3), Transferrin (Transferrin Human ELISA Kit, Invitrogen cat # EHTF), Hepcidin (Human Hepcidin Quantikine ELISA kit, R&D systems cat # DHP350) and Ferritin (Ferritin Human ELISA Kit, Invitrogen cat # EHFTL). Plasma samples were diluted 1:2 for EPO, 1: 20,000 for Transferrin, 1:5 for Hepcidin, and 1:10 for Ferritin using the kit manufacturer’s diluent. ELISAs were performed using the manufacturer’s protocol and absorbance was measured at 450 nm using the Synergy H4 plate reader. GraphPad Prism (Prism9) was used for statistical analysis and graphing data. Two-way ANOVA was used to determine p-values. A p-value of <0.05 was deemed significant.

## Results

### Subcohort of patients with multimorbid conditions at elevated risk of CVD

The OPOS sub-cohort analyzed herein consisted of 95 participants recruited for the OPOS study (358 participants). Participants with at least one condition: anemia, dyslipidemia, hypertension, T2DM and/or diagnosed HFpEF, were sampled as a sub-cohort. The demographics and clinical parameters of the OPOS cohort are shown in Table 3 and Supplemental Table 2. Some multimorbid parameters overlap between controls and the cases in each condition but otherwise served provided sex and age matched controls with CVD but without the specified outcome. Among these cases, dyslipidemia were associated with a higher NYHA score. BMI was higher in patients with T2DM and anemia but lower in those with HFpEF. Males were more prevalent in T2DM, hypertension, and dyslipidemia sub-populations. In contrast, females were more prevalent in anemia (taking into consideration the sex-dependent normal ranges used for males/females of the study population). Overall, this subcohort represents patients with salient risk factors requiring an intervention by cardiac surgery.

**Table 3.** Demographics of the OPOS sub-cohort.

|  | Total | Anemia |  |  | Dyslipidemia |  |  | Hypertension |  |  |
| --- | --- | --- | --- | --- | --- | --- | --- | --- | --- | --- |
|  | (n=95) | Cases<br>(n=31) | Controls<br>(n=64) | p-value | Cases<br>(n=66) | Controls<br>(n=29) | p-value | Cases<br>(n=70) | Controls<br>(n=25) | p-value |
| Age (years) | 63.5 ± 6.6 | 65.1 ± 5.8 | 62.7 ± 6.8 | 0.5 | 64.3 ± 6.3 | 61.7 ± 6.8 | 0.03 | 64.2 ± 6.9 | 61.6 ± 5.3 | **0.04** |
| Females | 30 (32%) | 16 (52%) | 14 (22%) |  | 17 (26%) | 13 (45%) |  | 17 (24%) | 13 (52%) |  |
| BMI (kg/m<sup>2</sup>) | 31.8 ± 7.3 | **34.4 ± 9.4** | **30.5 ± 5.6** | **0.007** | 31.3 ± 6.0 | 32.7 ± 9.6 | 0.70 | 31.4 ± 6.3 | 32.8 ± 9.6 | 0.10 |
| Hemoglobin (g/L) | 134.8 ± 14.7 | **117.8 ± 11.5** | **142.6 ± 7.8** | **<0.0001** | 134.8 ± 14.6 | 135.0 ± 14.8 | 0.60 | 134.9 ± 15.2 | 134.7 ± 12.9 | 0.82 |
| Hematocrit (L/L) | 0.40 ± 0.04 | **0.35 ± 0.03** | **0.42 ± 0.02** | **<0.0001** | 0.40 ± 0.04 | 0.40 ± 0.04 | 0.46 | 0.40 ± 0.04 | 0.40 ± 0.03 | 0.75 |
| MCV (fl) | 90.1 ± 4.3 | 88.2 ± 4.7 | 90.9 ± 3.8 | 0.052 | 89.4 ± 4.2 | 91.9 ± 3.9 | 0.054 | 90.3 ± 4.3 | 89.6 ± 4.2 | 0.55 |
| RBC (x10<sup>12</sup>/L) | 4.4 ± 0.4 | **4.0 ± 0.4** | **4.6 ± 0.3** | **<0.0001** | 4.5 ± 0.4 | 4.4 ± 0.4 | 0.81 | 4.4 ± 0.5 | 4.5 ± 0.3 | 0.53 |
| HbA1c (%) | 6.1 ± 1.0 | 6.2 ± 1.3 | 6.0 ± 0.9 | 0.68 | 6.2 ± 1.1 | 5.8 ± 0.8 | 0.29 | 6.1 ± 1.1 | 5.8 ± 0.8 | 0.42 |
| Cholesterol (mmol/L) | 4.1 ± 1.2 | 4.0 ± 1.1 | 4.1 ± 1.2 | 0.86 | 3.9 ± 1.1 | 4.5 ± 1.1 | 0.06 | 3.9 ± 1.2 | 4.4 ± 1.1 | 0.52 |
| HDL (mmol/L) | 1.2 ± 0.4 | 1.2 ± 0.4 | 1.2 ± 0.3 | 0.66 | 1.2 ± 0.3 | 1.4 ± 0.3 | 0.057 | 1.2 ± 0.4 | 1.3 ± 0.3 | 0.26 |
| LDL (mmol/L) | 2.1 ± 0.9 | 2.1 ± 0.9 | 2.1 ± 0.9 | 0.89 | **2.0 ± 0.8** | **2.5 ± 1.0** | **0.03** | 2.0 ± 0.9 | 2.5 ± 1.0 | 0.51 |
| LDL/HDL | 1.8 ± 0.8 | 1.7 ± 0.8 | 1.8 ± 0.7 | 0.63 | 1.7 ± 0.7 | 1.9 ± 0.7 | 0.38 | 1.7 ± 0.7 | 1.9 ± 0.8 | 0.92 |
| Non-HDL (mmol/L) | 2.9 ± 1.1 | 2.8 ± 1.0 | 2.9 ± 1.1 | 0.75 | 2.7 ± 1.1 | 3.1 ± 1.0 | 0.17 | 2.8 ± 1.1 | 3.1 ± 1.0 | 0.79 |
| Triglycerides (mmol/L) | 1.6 ± 1.1 | 1.8 ± 1.2 | 1.6 ± 1.0 | 0.58 | 1.7 ± 1.2 | 1.3 ± 0.6 | 0.27 | 1.7 ± 1.2 | 1.4 ± 0.6 | 0.59 |
| NLR | 3.9 ± 5.1 | 6.1 ± 8.5 | 2.9 ± 1.2 | 0.11 | 3.8 ± 4.6 | 4.1 ± 6.0 | 0.47 | 4.2 ± 5.7 | 3.1 ± 2.1 | 0.55 |

BMI=Body mass index, HDL=high-density lipoprotein, LDL=low-density lipoprotein, MCV=Mean Corpuscular Volume, RBC=red blood cells (erythrocytes), NLR=Neutrophils Lymphocytes ratio. p-value for continuous variable (BMI, Hemoglobin, Hematocrit, HbA1c, Cholesterol, HDL, LDL, MCV, Triglycerides, RBC, NLR) expressed as mean ±SD, p-value calculated using t-test. Categorical variable (Sex) expressed as number of cases (percentage of group), p-value calculated using Chi-square test. Significant p-value of <0.05 are bolded.

### Reassurance of experimental approach for avoiding SNP linkage disequilibrium

SNPs rs1617640(promoter), rs507392(intron) and rs551238(downstream) are located within 4.2 kb of one another. Due to this proximity, linkage disequilibrium is likely to occur[34]. Here, we performed a linkage disequilibrium analysis using SNPstats and found a high degree of linkage disequilibrium between our three SNPs (Supplemental Figure 1a). The highest degree of linkage disequilibrium was measured between rs1617640(promoter) and rs507392(intron), followed by rs507392(intron) and rs551238(downstream). SNPs rs1617640(promoter) and rs551238(downstream) were observed to have the least degree of disequilibrium. These associations correlated with the SNPs’ positions along the *EPO* gene, with those closest in proximity (Supplemental Figure 1b) showing the highest degree of disequilibrium.

### The presence of the C allele in rs1617640(promoter) increases the risk of anemia

Each SNP was examined for association based on dominant, recessive, or additive inheritance models (Table 4). The C allele of rs1617640 SNP in the promoter region was associated with anemia in an additive inheritance model with an odds ratio (OR) of 1.96 (p-value 0.04). Whether clinical parameters correlated with specific genotypes was then investigated. In SNP rs1617640, there were significant differences between genotypes in hemoglobin, neutrophil to lymphocyte ratios (NLR), cholesterol, low-density lipoprotein (LDL), and non-high-density lipoprotein (non-HDL) (Supplemental Table 3, Supplemental Figure 2). The lower hemoglobin levels in the CC genotype support the association of anemia with the C allele, with the higher NLR suggesting elevated inflammation. The rs1617640(promoter) CC genotype was associated with a lower plasma EPO level, although iron bioavailability (as measured by Ferritin, Hepcidin and Transferrin) was not significantly different (Figure 1). These findings suggest that SNP rs1617640(promoter) is likely affecting EPO directly, not through iron bioavailability.

**Figure 1.**
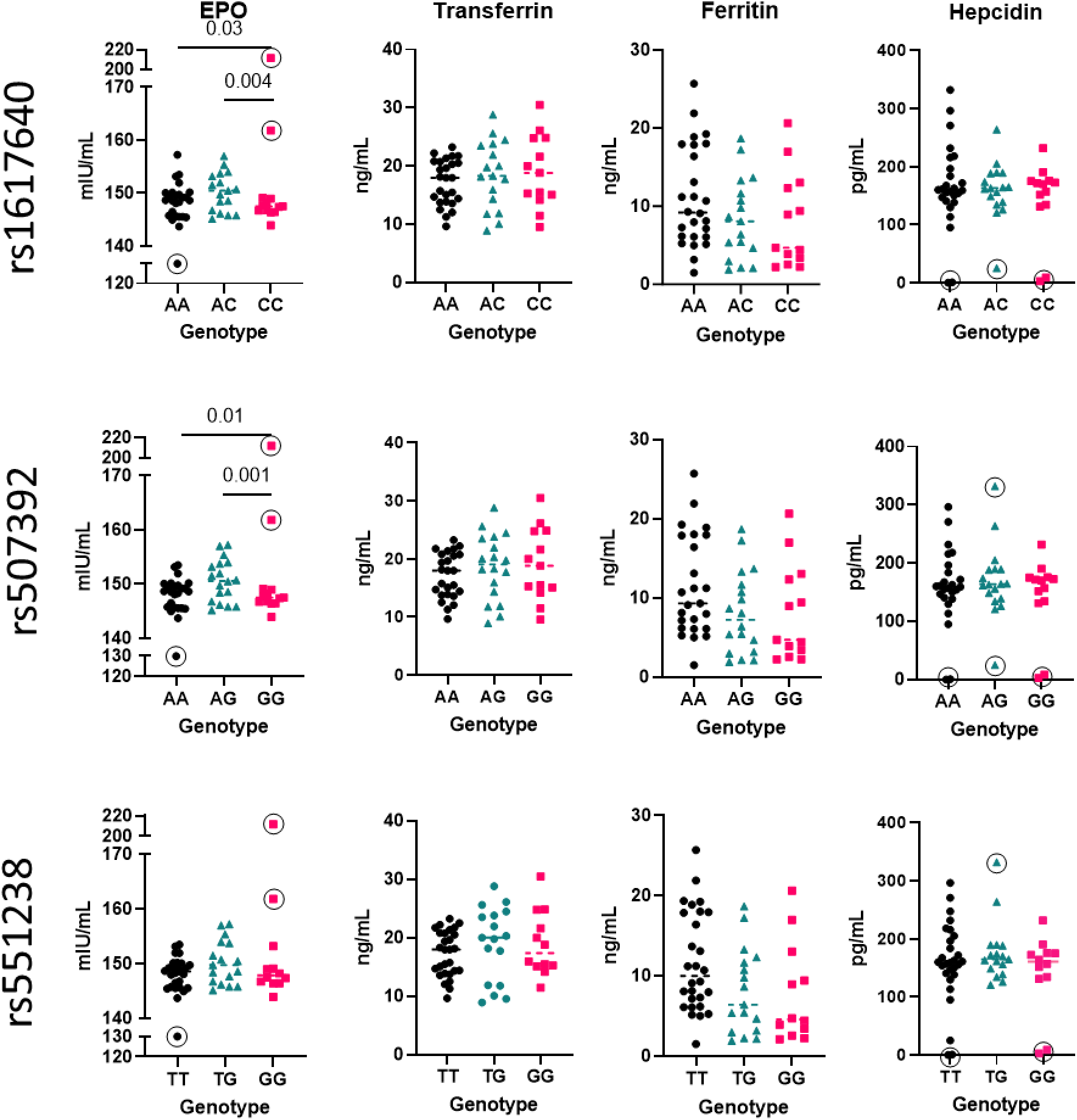
Plasma EPO concentration is decreased in rs161640 CC genotype and rs507392 GG genotype, but not rs51238. Transferrin and Hepcidin showed no genotype correlation although there does appear to be variation in Ferritin it does not reach significance. Plasma concentrations were determined using ELISA assays with outliers marked by circles and excluded from statistical analysis. Outliers were determined by ROUT test and *a priori* as two standard deviations from the mean or else being below the lower limit of detection of the ELISA. ANOVA was used to determine significance with p-values < 0.05 deemed significant.

**Table 4.**
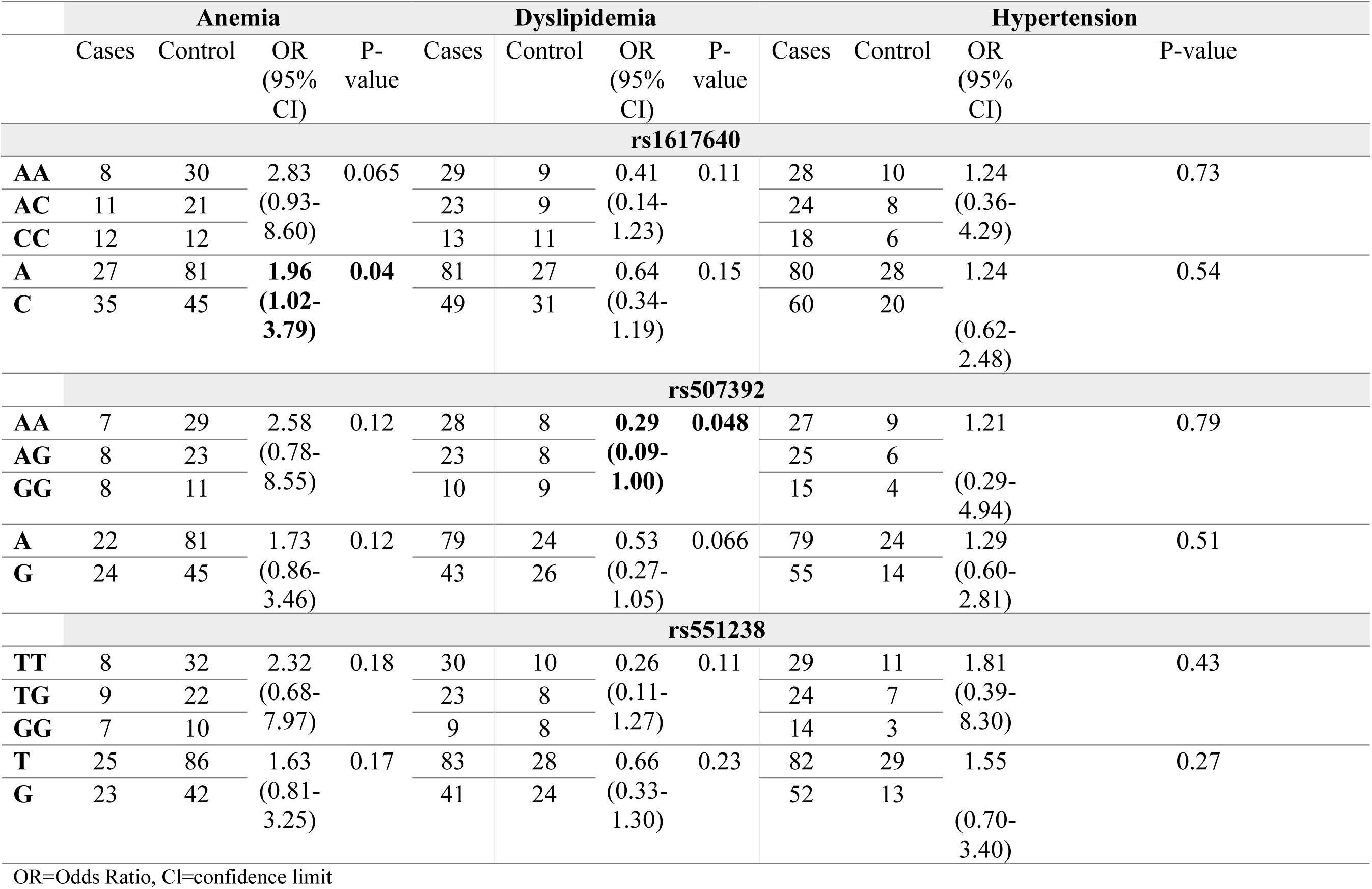
Association of anemia, dyslipidemia and hypertension by genotype.

### GG Genotype of rs507392(intron) is protective against dyslipidemia

To characterize the role of the GG Genotype of intron SNP (rs507392) we examined the OR-value for anemia, dyslipidemia, and hypertension. The GG genotype of rs507392 was associated with protection against dyslipidemia in a recessive model (OR value = 0.29; p-value = 0.048) (Table 4), while there was no significance with anemia or hypertension. Variations based on genotype in rs507392(intron) were seen in clinical parameters including cholesterol, LDL, non-HDL, and NLR (Supplemental Table 4, Supplemental Figure 3). This suggests that SNP rs507392(intron) is associated with lipid handling/regulation. As with rs1617640(promoter), the GG genotype in rs507392(intron) demonstrated lower levels of EPO in the plasma (Figure 1). There was no association to risk factors for SNP rs551238(downstream) (Table 4), and fewer significant differences in clinical parameters were observed (Supplemental Table 5 and Supplemental Figure 4). Unlike rs1617640(promoter) and rs507392(intron), no significant EPO changes in rs551238(downstream) were observed (Figure 1).

### No strong evidence for EPO haplotype association with risk factors

Haplotype was also examined to determine the cumulative effects of all three SNPs to clinical outcomes. The predominant haplotypes of AAT and CGG did not correlate with any clinical phenotype. However, one of the rare haplotypes of CGT (which occurred in approximately 3% of our cohort) did correlate with dyslipidemia. The OR-value of 0.05 suggests that the CGT haplotype is protective against dyslipidemia but should be interpreted with caution given the low prevalence (Table 5, Supplemental Table 6).

**Table 5.**
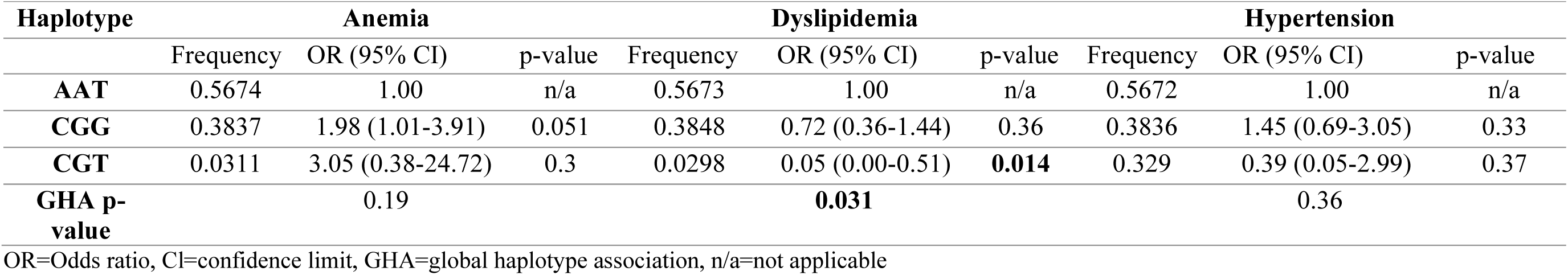
Haplotype association.

| Haplotype | Anemia |  |  | Dyslipidemia |  |  | Hypertension |  |  |
| --- | --- | --- | --- | --- | --- | --- | --- | --- | --- |
|  | Frequency | OR (95% CI) | p-value | Frequency | OR (95% CI) | p-value | Frequency | OR (95% CI) | p-value |
| **AAT** | 0.5674 | 1.00 | n/a | 0.5673 | 1.00 | n/a | 0.5672 | 1.00 | n/a |
| **CGG** | 0.3837 | 1.98 (1.01-3.91) | 0.051 | 0.3848 | 0.72 (0.36-1.44) | 0.36 | 0.3836 | 1.45 (0.69-3.05) | 0.33 |
| **CGT** | 0.0311 | 3.05 (0.38-24.72) | 0.3 | 0.0298 | 0.05 (0.00-0.51) | **0.014** | 0.329 | 0.39 (0.05-2.99) | 0.37 |
| **GHA p-value** |  | 0.19 |  |  | **0.031** |  |  | 0.36 |  |

OR=Odds ratio, CI=confidence limit, GHA=global haplotype association, n/a=not applicable

### Sex differences are apparent with SNP associations to risk factors

Precision medicine necessitates accounting for sex differences, and here we find meaningful sex effects to the associated risk factors. In males, the prevalence of dyslipidemia, T2DM, and hypertension are significantly higher than that observed in females[35], whereas in females, the prevalence of anemia[36] and HFpEF[37] is significantly higher. Given these sex differences in the clinical phenotypes examined herein, we analyzed the differences in SNPs within each sex to determine if sex differences would affect our results. SNP rs507392(intron) AG and GG genotypes, and rs551238(downstream) TG genotype, were associated with hypertension in females, but not in males (Table 6, Supplemental Table 7), suggesting that the rs507392(intron) and rs551238(downstream) SNPs could be risk factors for hypertension in females but not males.

**Table 6.**
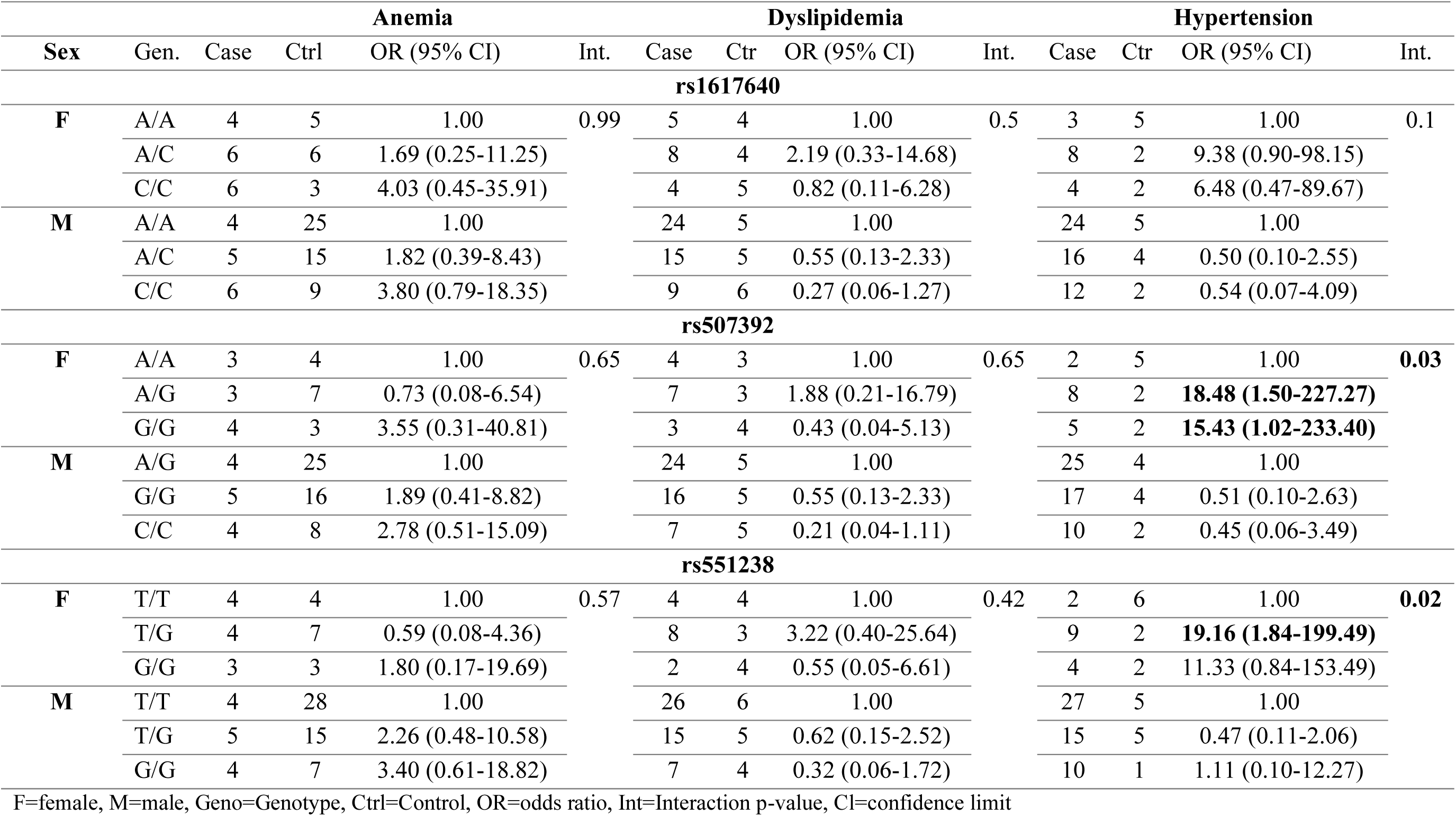
Sex differences.

### New EPO variants of potential concern identified

During the analysis of *a priori* sequences obtained for rs1617640(promoter), rs507392(intron), and rs551238(downstream), other variants were also identified (Supplemental Table 8). Many of these variants were only detected in a single individual as a heterozygote; others were identified in a several individuals, with most being heterozygotes. However, most of these variants have not previously been reported and are, therefore, of unknown significance. Three variants were found in a significant subset of our population, including a four-base pair deletion (2167?-2170?del). This variant is located within a repeat region in intron 3, and thus the breakpoint may be inexact. This base pair deletion was found to be homozygous in 76 cases and suspected to be heterozygous in 4 cases. The 3154T>G substitution was found in 83 individuals, with 77 of these determined as homozygotes. Lastly, 3806T>C was found in 33 individuals, with 31 of these determined as heterozygotes. This data suggests more variation in EPO than previously reported, which needs further exploration in larger study populations where further associations can be uncovered.

## Discussion

Here, we identified linkage between the three *EPO* SNPs and cardiovascular disease risk factors, including dyslipidemia and HFpEF-associated comorbidities: anemia, T2DM, and hypertension. The CC genotype of rs1617640(promoter) and GG genotype of rs507392(intron) were associated with lower EPO plasma levels and correlated with comparably higher cholesterol, LDL, and non-HDL lipoproteins. The promoter-SNP (rs1617640) CC was associated with lower Hb and anemia in a log additive model and has been previously linked to anemia in T2DM[19]. While the role of the promoter-SNP in CVD has been explored to a lesser extent, our data support it as being linked to anemia in CVD. Surprisingly, the intron-SNP (s507392) GG genotype had a protective association with dyslipidemia in a recessive model. There was no significant association between hypertension and the SNPs in the overall population, although rs507392(intron) and rs551238(downstream) were associated with hypertension in females, suggesting a sex dependence. Our study highlights that EPO variants are associated with comorbid conditions that present significant cardiovascular risk factors of anemia, dyslipidemia, and hypertension for the first time.

EPO levels are well known to be associated with anemia[23,24], so any SNP that alters plasma EPO levels, such as rs1617640(promoter) and rs507392(intron), as we have shown, could be risk factors for anemia, and require additional pharmacogenetic guided intervention. Yet, iron bioavailability also contributes to two of the most common forms, iron-deficiency anemia and anemia of inflammation (or anemia of chronic disease, e.g., CVD)[38]. The rs1617640(promoter) C-allele associated with anemia was also associated with NLR, indicative of active inflammatory processes[39,40]. This suggests the C allele for rs1617640(promoter) is associated with inflammation and decreased Hb. It has been purported that EPO can exert an indirect anti-oxidant effect by utilizing iron in erythrocytosis resulting in less bioavailable iron.[41] Patients with anemia of inflammation commonly have reduced serum iron and normal to elevated iron stores.[42] In blood, iron is transported by transferrin[43] and can be stored in ferritin within most cells, like macrophages and hepatocytes.[44] The release of recycled iron from intracellular stores is partially regulated by hepcidin—a hormone that inhibits the transportation of intracellular iron to the blood and is induced by inflammation.[38] Hypoxia-inducible factors (HIFs; which upregulate *EPO*[7]) also reduce hepcidin, thus increasing transferrin, ferritin, and iron bioavailability.[45] We measured hepcidin, transferrin and ferritin to examine whether the *EPO* SNPs altered iron bioavailability. Still, despite elevated NLR associated with allelic frequency, we did not find significant differences between genotypes. This leads us to conclude that the SNPs in *EPO* are likely not exerting their effects by regulating iron bioavailability.

Ours is the first study to identify *EPO* SNPs associated with dyslipidemia and/or lipoproteins in medically managed patients. Prior studies have reported that rhEPO therapy modulates triglycerides, cholesterol, and other lipoproteins [26–28]. In a mouse model of CKD, treatment with rhEPO resulted in decreased triglycerides, cholesterol and LDL, but not HDL[26]. In the present study, the GG genotype of rs507392(intron) correlated with lower EPO concentrations but increased cholesterol, LDL and non-HDL. These variations would also be expected with diagnosed dyslipidemia, yet the GG genotype was associated with resistance to dyslipidemia. This apparent contradiction is, however, more likely a consequence of medical management of dyslipidemia (e.g., statins) to lower lipoprotein levels[46], not the association to the clinical outcome. Future studies could be designed to validate this epidemiologically with sufficient covariant adjustments. While rs1617640(promoter) and rs551238(downstream) did not associate with dyslipidemia as an outcome, there was significant variation in cholesterol, with the rs1617640(promoter)AC variant genotype being significantly lower for LDL and non-HDL lipoproteins. The rs551238(downstream) TG variant was significantly associated with lower cholesterol and non-HDL. The CGT haplotype was found to be protective against dyslipidemia, but this was a rare finding (less than 3% of cohort) and should be interpreted with caution. Together, these results show that *EPO* SNPs influence lipid metabolism and that rs507392(intron) is associated with dyslipidemia. The fundamental biology and molecular biology of EPO in these processes need to be explored further.

Many clinical phenotypes, including those examined here, have sex differences attributable to prevalence, response to treatment, or clinical outcomes. Hypertension did not show an association with the SNPs with the sexes combined, yet the presence of the G allele in rs507392(intron) and the TG genotype in rs551238(downstream) showed an elevated risk in females but not males. Hypertension is more prevalent in males[35], and our cohort contained more males than females, which might explain why this association was initially masked. The relationship between estrogen and the renin-angiotensin-aldosterone system (RAAS) is thought to be one of the main reasons for the sex differences observed in hypertension. The RAAS has also been found to be modulated by EPO: in Wistar rats, rhEPO increases renin[47] and in isolated rat kidneys, rhEPO promotes angiotensin II (AngII) production.[48] In the oviduct and endometrium, estrogen (17β (E2)) can also promote the expression of *EPO*.[49,50] Given the relationship between EPO and the RAAS system by estrogen, this could be a potential mechanism for why the G allele of rs507392 and TG genotype of rs551238 are risk factors for hypertension just in females. Although this finding does require validation in a larger cohort (including more females with a direct assessment of sex hormones), this may be a significant finding since hypertension tends to be a more salient risk factor for CVD in females.[51]

Our results suggest that *EPO* SNPs may be associated with risk factors and co-morbidities of CVD by affecting the amount of EPO produced. Given the ability to modulate the amount of EPO either directly through rhEPO or indirectly by targeting upstream of EPO using hypoxia-inducible factor prolyl-hydroxylase inhibitors, these SNPs could be used to inform the best course of medical management. Hypoxia-inducible factor (HIF) prolyl-hydroxylase inhibitors increase EPO by stabilizing the HIFs that transcriptionally act upon *EPO* DNA under normoxic conditions.[52,53] Though not yet established, should *EPO* SNP result in less HIF binding to *EPO* in transcriptional regulation or stability of the intronic stores of EPO mRNA[54], then the use of HIF prolyl-hydroxylase inhibitors may be less potent for directly increasing EPO compared to rhEPO. Although more work is needed before these SNPs are applied clinically, they could guide pharmacogenetic interventions that seek to increase EPO or address anemia in the future or reclassify existing clinical trial data for subgroup analyses. In addition to the principally known SNPs, we also identified novel variants. Most occurred as heterozygotes in a single individual, but four occurred in multiple individuals and are potential polymorphisms. In clinical care, sequencing is more commonplace because of the integration of next-generation sequencing technologies into standard care. In other disorders, this led to increased variant detection.[55] Surprisingly, *EPO* has long been overlooked on many gene arrays or panels, which may account for the low prevalence of *EPO* variants in the literature. As the sequencing of *EPO* increases, we anticipate further variants of concern can be detected, with a potential for linkages in large epidemiology studies now that we have discovered them.

Our study is not without limitations. One was the small sample size and depth of patient file analyses, which did not allow us to account for many covariates (such as ethnicity, other comorbidities, or medical management) that could affect our results. Our study did identify dyslipidemia as a novel disorder and can direct further focused sequencing toward cases of dyslipidemia and potentially investigate use/efficacy of statins as a covariant. Additionally, hypertension was identified in females but not males, with the rs507392 G allele and rs551238 TG genotype being risk factors. As we had relatively few females, more sequencing should target females with hypertension. Even with our sample size, we have identified variables that can benefit epidemiological or database studies to validate clinical relevance, screening risk or precision medical management.

Here, we have established that *EPO* SNP rs1617640 C-allele was associated with anemia, as well as changes in lipid metabolism and inflammation. We show the association of GG genotype for rs507392 to dyslipidemia for the first time. This genotype indicates effects on lipid profiles despite standard care. In females only, we show that the rs507392 and rs551238 *EPO* SNPs are associated with hypertension. Together, our results suggest that SNPs in *EPO* are involved in Hb regulation, lipid metabolism, and inflammation with a female-specific effect on blood pressure.

## Data Availability

All data produced in the present study are available upon reasonable request to the authors

## Acknowledgements

Blood samples for gDNA and clinical data were obtained from the IMPART BioBank (https://impart.team/biobank/). Clinical data and samples were retrieved by IMPART BioBank staff, including Dr. Christie Aguiar, Angella Mercer, and Saumil Shah, in collaboration with the authors who extend their appreciation. The IMPART BioBank was supported by endowed funds from the Dalhousie Medical Research Foundation and specialized equipment provided by Saint John Regional Hospital Foundation-directed donation. This study was supported by the Natural Sciences and Engineering Research Council (NSERC) [discovery grant #RGPIN-2018-05626 and # RGPIN-(2020)-04878] (JAS & KRB), Heart & Stroke Foundation of New Brunswick and the generosity of the Chesley Family Grant (KRB). VN was supported by the NSERC Alexander Graham Bell Canada Graduate Scholarship-Doctoral (CGS-D) and the New Brunswick Health Research Foundation Beatrice Hunter Cancer Research Training Program Award.

## Authorship Contributions

Conceived; designed experimental approaches of the study (VN, JAS, KRB); Mentored trainee acquisition of data and execution of the study (JAS & KRB); Collected and assembled data for the study (VN, ALE, VLN, KD, AH, TP, PK, JFL, & KRB); Analyzed; interpreted & reported data (VN, AE, VLN, & KRB); Drafted manuscript (VN & KRB); critically reviewed, revised and edited manuscript for intellectual content and approved final work (VN, ALE, VLN, KD, JFL, JAS, & KRB).

