## Supplemental Figures for "Erythropoietin single nucleotide polymorphisms are associated with anemia and dyslipidemia in a cardiovascular disease population"

^6^IMPART investigator team Canada (https://impart.team/)

**Supplemental Material**

**Supplemental Table 1- Erythropoietin SNP associations**

| SNP | Allele | Phenotype | References |
| --- | --- | --- | --- |
| rs1617640 | G | Higher Hct in healthy blood donors | ^1^ |
|  |  | Diabetic microvascular complications | ^2–10^ |
|  |  | Decreased overall mortality in diabetes | ^11^ |
|  |  | Higher Hb, Hct, RBC count and earlier onset PAD | ^12^ |
|  |  | MDS and ALL | ^13^ |
|  |  | Improved cognitive performance in Schizophrenia | ^14^ |
|  | T | Anemia in T2DM | ^15^ |
|  |  | Renal dysfunction following cardiac surgery | ^16^ |
|  |  | Improved response to platinum-based chemotherapy in NSCLC | ^17^ |
| rs507392 | C | Diabetic microvascular complications | ^2,4,5,18^ |
|  |  | Decreased overall mortality in diabetes | ^11^ |
| rs551238 | C | Higher Hct in healthy blood donors | ^1^ |
|  |  | Diabetic microvascular complications | ^1,2,4–7,18,19^ |
|  |  | Pre-term infant brain injury | ^20^ |

SNP=Single Nucleotide Polymorphism, Hct=hematocrit, Hb=hemoglobin, RBC=red blood cell, PAD=peripheral artery disease, MDS=myelodysplatic syndrome, ALL=acute lymphoblastic leukemia, T2DM=type 2 diabetes mellites, NSCLC=non-small cell lung cancer

**Supplemental Table 2 - Demographics and clinical parameters of the OPOS cohort by disorder**

|  | **Total** | **Anemia** | | | **Dyslipidemia** | | | **Hypertension** | | |
| --- | --- | --- | --- | --- | --- | --- | --- | --- | --- | --- |
|  | (n=95) | Cases (n=31) | Controls (n=64) | p-value | Cases (n=66) | Controls (n=29) | p-value | Cases (n=70) | Controls (n=25) | p-value |
| Age (years) | 63.5 ± 6.6 | 65.1 ± 5.8 | 62.7 ± 6.8 | 0.52 | 64.3 ± 6.3 | 61.7 ± 6.8 | **0.03** | 64.2 ± 6.9 | 61.6 ± 5.3 | **0.04** |
| Males | 65 (68%) | 15 (48%) | 50 (78%) | **0.0035** | 49 (74%) | 16 (55%) | **0.045** | 53 (76%) | 12 (48%) | **0.01** |
| Females | 30 (32%) | 16 (52%) | 14 (22%) |  | 17 (26%) | 13 (45%) |  | 17 (24%) | 13 (52%) |  |
| BMI (kg/m^2^) | 31.8 ± 7.3 | 34.4 ± 9.4 | 30.5± 5.6 | **0.007** | 31.3 ± 6.0 | 32.7 ± 9.6 | 0.70 | 31.4 ± 6.3 | 32.8 ± 9.6 | 0.10 |
| Normal | 14 (15%) | 4 (13%) | 10 (16%) | 0.19 | 9 (14%) | 5 (17%) | 0.86 | 11 (16%) | 3 (12%) | 0.83 |
| Pre-obese | 31 (33%) | 8 (26%) | 23 (36%) |  | 22 (33%) | 9 (31 %) |  | 22 (31%) | 9 (36%) |  |
| Obese 1 | 22 (23%) | 6 (19%) | 16 (25%) |  | 17 (26%) | 5 (17%) |  | 16 (23%) | 6 (24%) |  |
| Obese 2 | 13 (14%) | 4 (13%) | 9 (14%) |  | 8 (12%) | 5 (17%) |  | 11 (16%) | 2 (8%) |  |
| Obese 3 | 15 (16%) | 9 (29%) | 6 (9%) |  | 10 (15%) | 5 (17%) |  | 10 (14%) | 5 (20%) |  |
| NYHA 1 | 18 (19%) | 4 (14%) | 14 (22%) | 0.42 | 8 (12%) | 10 (37%) | **0.006** | 13 (19%) | 5 (22%) | 0.91 |
| 2 | 39 (42%) | 11 (38%) | 28 (44%) |  | 31 (47%) | 8 (30%) |  | 29 (41%) | 10 (43%) |  |
| 3 | 25 (27%) | 11 (38%) | 14 (22%) |  | 16 (24%) | 9 (33%) |  | 19 (27%) | 6 (26%) |  |
| 4 | 11 (12%) | 3 (10%) | 8 (13%) |  | 11 (17%) | 0 (0%) |  | 9 (13%) | 2 (9%) |  |
| LVEF (%) | 59.8 ± 11.2 | 60.9 ± 10.7 | 59.3 ± 11.4 | 0.99 | 58.0 ± 11.9 | 64.0 ± 7.8 | 0.07 | 60.5 ± 11.3 | 58.1 ± 10.7 | 0.26 |
| LVEDP (mmHg) | 18.0 ± 6.6 | 18.8 ± 6.4 | 17.7 ± 6.7 | 0.052 | 18.6 ± 6.7 | 16.6 ± 6.4 | 0.10 | 18.7 ± 7.0 | 15.7 ± 4.8 | 0.10 |
| Hemoglobin (g/L) | 134.8 ± 14.7 | 117.8 ± 11.5 | 142.6 ± 7.8 | **<0.0001** | 134.8 ± 14.6 | 135.0 ± 14.8 | 0.60 | 134.9 ± 15.2 | 134.7 ± 12.9 | 0.82 |
| Hematocrit (L/L) | 0.40 ± 0.04 | 0.35 ± 0.03 | 0.42 ± 0.02 | **<0.0001** | 0.40 ± 0.04 | 0.40 ± 0.04 | 0.46 | 0.40 ± 0.04 | 0.40 ± 0.03 | 0.75 |
| MCV(fL) | 90.1 ± 4.3 | 88.2 ± 4.7 | 90.9 ± 3.8 | 0.052 | 89.4 ± 4.2 | 91.9 ± 3.9 | 0.054 | 90.3 ± 4.3 | 89.6 ± 4.2 | 0.55 |
| HbA1c (%) | 6.1 ± 1.0 | 6.2 ± 1.3 | 6.0 ± 0.9 | 0.68 | 6.2 ± 1.1 | 5.8 ± 0.8 | 0.29 | 6.1 ± 1.1 | 5.8 ± 0.8 | 0.42 |
| Chol (mmol/L) | 4.1 ± 1.2 | 4.0 ± 1.1 | 4.1 ± 1.2 | 0.86 | 3.9 ± 1.1 | 4.5 ± 1.1 | 0.07 | 3.9 ± 1.2 | 4.4 ± 1.1 | 0.52 |
| HDL (mmol/L) | 1.2 ± 0.4 | 1.2 ± 0.4 | 1.2 ± 0.3 | 0.66 | 1.2 ± 0.3 | 1.4 ± 0.3 | 0.057 | 1.2 ± 0.4 | 1.3 ± 0.3 | 0.26 |
| LDL (mmol/L) | 2.1 ± 0.9 | 2.1 ± 0.9 | 2.1 ± 0.9 | 0.89 | 2.0 ± 0.8 | 2.5 ± 1.0 | **0.03** | 2.0 ± 0.9 | 2.5 ± 1.0 | 0.51 |
| LDL/HDL | 1.8 ± 0.8 | 1.7 ± 0.8 | 1.8 ± 0.7 | 0.63 | 1.7 ± 0.7 | 1.9 ± 0.7 | 0.38 | 1.7 ± 0.7 | 1.9 ± 0.8 | 0.92 |
| Non-HDL (mmol/L) | 2.9 ± 1.1 | 2.8 ± 1.0 | 2.9 ± 1.1 | 0.75 | 2.7 ±1.1 | 3.1 ± 1.0 | 0.17 | 2.8 ± 1.1 | 3.1 ± 1.0 | 0.79 |
| Trigly (mmol/L) | 1.6 ± 1.1 | 1.8 ± 1.2 | 1.6 ± 1.0 | 0.58 | 1.7 ± 1.2 | 1.3 ± 0.6 | 0.27 | 1.7 ± 1.2 | 1.4 ± 0.6 | 0.59 |
| RBC (x10^12/L) | 4.4 ± 0.4 | 4.0 ± 0.4 | 4.6 ± 0.3 | **<0.0001** | 4.5 ± 0.4 | 4.4 ± 0.4 | 0.81 | 4.4 ± 0.5 | 4.5 ± 0.3 | 0.53 |
| WBC (x10^9/L) | 7.4 ± 2.4 | 7.8 ± 3.2 | 7.2 ± 1.9 | 0.37 | 7.6 ± 2.6 | 6.8 ± 1.8 | 0.44 | 7.4 ± 2.4 | 7.3 ± 2.5 | 0.46 |
| Platlets (x10^9/L) | 205.5 ± 53.9 | 209.0 ± 72.2 | 203.9 ± 43.1 | 0.56 | 209.5 ± 57.1 | 195.8 ± 43.7 | 0.38 | 205.7 ± 56.5 | 204.9 ± 45.4 | 0.91 |
| Neutro (x10^9/L) | 4.9 ± 2.2 | 5.5 ± 3.3 | 4.7 ± 1.5 | 0.21 | 5.1 ± 2.3 | 4.5 ± 1.8 | 0.56 | 5.0 ± 2.2 | 4.8 ± 2.2 | 0.62 |
| Lympho (x10^9/L) | 1.7 ± 0.6 | 1.5 ± 0.7 | 1.7 ± 0.6 | 0.23 | 1.7 ± 0.6 | 1.6 ± 0.6 | 0.99 | 1.6 ± 0.7 | 1.7 ± 0.6 | 0.20 |
| NLR | 3.9 ± 5.1 | 6.1 ± 8.5 | 2.9 ± 1.2 | 0.11 | 3.8 ± 4.6 | 4.1 ± 6.0 | 0.47 | 4.2 ± 5.7 | 3.1 ± 2.1 | 0.55 |
| Troponin (ng/L) | 785.2 ±590.3 | 878.6 ± 823.7 | 740.9 ± 431.0 | 0.61 | 694.1 ± 428.4 | 999.1 ± 818.6 | **0.03** | 794.3 ± 637.3 | 758.6 ± 420.7 | 0.51 |
| Creatine (mmol/L) | 92.9 ± 71.1 | 113.3 ± 121.6 | 83.7 ± 19.4 | 0.22 | 97.8 ± 82.6 | 81.1 ± 22.9 | 0.25 | 89.0 ± 26.4 | 105.1 ± 134.6 | 0.22 |
| Urea (mmol/L) | 6.7 ± 2.5 | 7.2 ± 3.3 | 6.5 ± 2.1 | 0.054 | 7.0 ± 2.7 | 6.1 ± 2.0 | 0.07 | 7.0 ± 2.3 | 5.8 ± 2.8 | 0.08 |

**Supplemental Table 2 cont’d - Demographics and clinical parameters of the OPOS cohort by disorder**

|  | **HFpEF** | | | **T2DM** | | |
| --- | --- | --- | --- | --- | --- | --- |
|  | Cases (n=77) | Controls (n=18) | p-value | Cases (n=33) | Controls (n=62) | p-value |
| Age (years) | 63.9 ± 6.8 | 61.9 ± 5.5 | 0.25 | 64.6 ± 5.5 | 63.0 ± 7.1 | 0.17 |
| Males | 53 (69%) | 12 (67%) | 0.86 | 26 (81%) | 36 (59%) | **0.040** |
| Females | 24 (31%) | 6 (33%) |  | 6 (19%) | 25 (41%) |  |
| BMI (kg/m^2^) | 30.9 ± 6.2 | 35.5 ± 10.0 | **0.016** | 33.3 ± 6.0 | 30.9 ± 7.8 | **0.010** |
| Normal | 12 (16%) | 2 (11%) | 0.11 | 1 (3%) | 13 (21%) | 0.14 |
| Pre-obese | 29 (38%) | 2 (11%) |  | 10 (30%) | 21 (34%) |  |
| Obese 1 | 16 (21%) | 6 (33%) |  | 10 (30%) | 12 (19%) |  |
| Obese 2 | 8 (10%) | 5 (38%) |  | 5 (15%) | 8 (13%) |  |
| Obese 3 | 12 (16%) | 3 (17 %) |  | 7 (21%) | 8 (13%) |  |
| NYHA 1 | 14 (18%) | 4 (25%) | 0.85 | 5 (15%) | 13 (22%) | 0.46 |
| 2 | 32 (42%) | 7 (44%) |  | 12 (36 %) | 27 (45%) |  |
| 3 | 22 (29%) | 3 (19%) |  | 12 (36%) | 13 (22%) |  |
| 4 | 9 (12%) | 2 (13%) |  | 4 (12%) | 7 (12%) |  |
| LVEF (%) | 63.9 ±7.4 | 42.6 ± 7.6 | **<0.0001** | 56.7 ± 11.3 | 61.5 ± 10.7 | 0.10 |
| LVEDP (mmHg) | 17.4 ± 6.5 | 20.4 ± 6.8 | 0.11 | 18.8 ± 6.6 | 17.6 ± 6.6 | 0.35 |
| Hemoglobin (g/L) | 135.0 ± 14.6 | 134.3 ± 14.9 | 0.86 | 132.9 ± 14.9 | 135.9 ± 14.4 | 0.24 |
| Hematocrit (L/L) | 0.40 ± 0.04 | 0.40 ± 0.04 | 0.71 | 0.39 ± 0.04 | 0.40 ± 0.04 | 0.22 |
| MCV(fL) | 90.1 ± 4.3 | 90.1 ± 3.9 | 0.98 | 89.5 ± 4.1 | 90.4 ± 4.3 | 0.22 |
| HbA1c (%) | 6.0 ± 1.0 | 6.4 ± 0.9 | 0.20 | 7.0 ± 1.2 | 5.6 ± 0.3 | **<0.0001** |
| Chol (mmol/L) | 4.1 ± 1.2 | 3.9 ± 1.1 | 0.62 | 3.6 ± 0.9 | 4.3 ± 1.2 | **0.012** |
| HDL (mmol/L) | 1.2 ± 0.4 | 1.2 ± 0.2 | 0.69 | 1.1 ± 0.4 | 1.3 ± 0.3 | **0.045** |
| LDL (mmol/L) | 2.1 ± 0.9 | 2.1 ± 1.1 | 0.84 | 1.7 ± 0.7 | 2.3 ± 1.0 | **0.016** |
| LDL/HDL | 1.8 ± 0.7 | 1.8 ± 0.9 | 0.70 | 1.6 ± 0.7 | 1.9 ± 0.7 | 0.60 |
| Non-HDL (mmol/L) | 2.9 ± 1.1 | 2.8 ± 1.1 | 0.82 | 2.5 ± 0.8 | 3.0 ± 1.1 | 0.06 |
| Trigly(mmol/L) | 1.6 ± 1.1 | 1.5 ± 0.7 | 0.54 | 1.9 ± 1.2 | 1.5 ± 0.9 | 0.71 |
| RBC (x10^12/L) | 4.5 ± 0.4 | 4.4 ± 0.5 | 0.63 | 4.4 ± 0.4 | 4.5 ± 0.4 | 0.58 |
| WBC (x10^9/L) | 7.3 ± 2.3 | 7.7 ± 2.9 | 0.57 | 8.0 ± 2.9 | 7.1 ± 2.0 | 0.26 |
| Platelets (x10^9/L) | 204.9 ± 56.6 | 208.1 ± 38.4 | 0.83 | 218.2 ± 68.9 | 198.5 ± 42.0 | 0.36 |
| Neutro (x10^9/L) | 4.9 ± 2.2 | 5.1 ± 2.5 | 0.75 | 5.5 ± 2.7 | 4.6 ± 1.8 | 0.15 |
| Lympho (x10^9/L) | 1.7 ± 0.7 | 1.8 ± 0.5 | 0.56 | 1.7 ± 0.7 | 1.7 ± 0.6 | 0.55 |
| NLR | 4.0 ± 5.6 | 3.1 ± 1.7 | 0.51 | 4.7 ± 6.1 | 3.5 ± 4.3 | 0.25 |
| Troponin (ng/L) | 838.1 ± 616.9 | 509.6 ± 300.8 | 0.057 | 754.1 ± 709.1 | 809.2 ± 520.8 | 0.42 |
| Creatine (mmol/L) | 85.7 ± 24.0 | 127.7 ± 158.5 | **0.032** | 110.8 ± 112.8 | 83.1 ± 23.6 | 0.25 |
| Urea (mmol/L) | 6.6 ± 2.1 | 7.2 ± 3.8 | 0.47 | 7.7 ± 3.1 | 6.2 ± 1.9 | **0.028** |

NYHA=New York Heart Association, BMI=Body mass index, HDL=high-density lipoprotein, LDL=low-density lipoprotein, LVEF=Left ventricular ejection fraction, LVEDP=left ventricular end diastolic pressure, HFpEF=Heart failure with preserved ejection fraction, T2DM=type 2 diabetes mellitus, REB=red blood cells (erythrocytes), WBC=white blood cells (leukocytes), Neutro=Neutrophiles, Lympho= Lymphocytes, NLR=Neutrophiles Lymphocytes ratio. p-value for continuous variable (BMI, LVEF, LVEDP, Hemoglobin, Hematocrit, HbA1c, Chol=Cholesterol, HDL, LDL, Trigly=Triglycerides, RBC, Troponin, Creatine, WBC, Neutrophiles, Lymphocytes, NLR, Urea) expressed as mean ±SD; p-value calculated using t-test. Categorical variable (Sex, BMI Class, and NYHA) expressed as number of cases (percentage of group); p-value calculated using Chi-square test. Significant p-values of <0.05 are bolded.

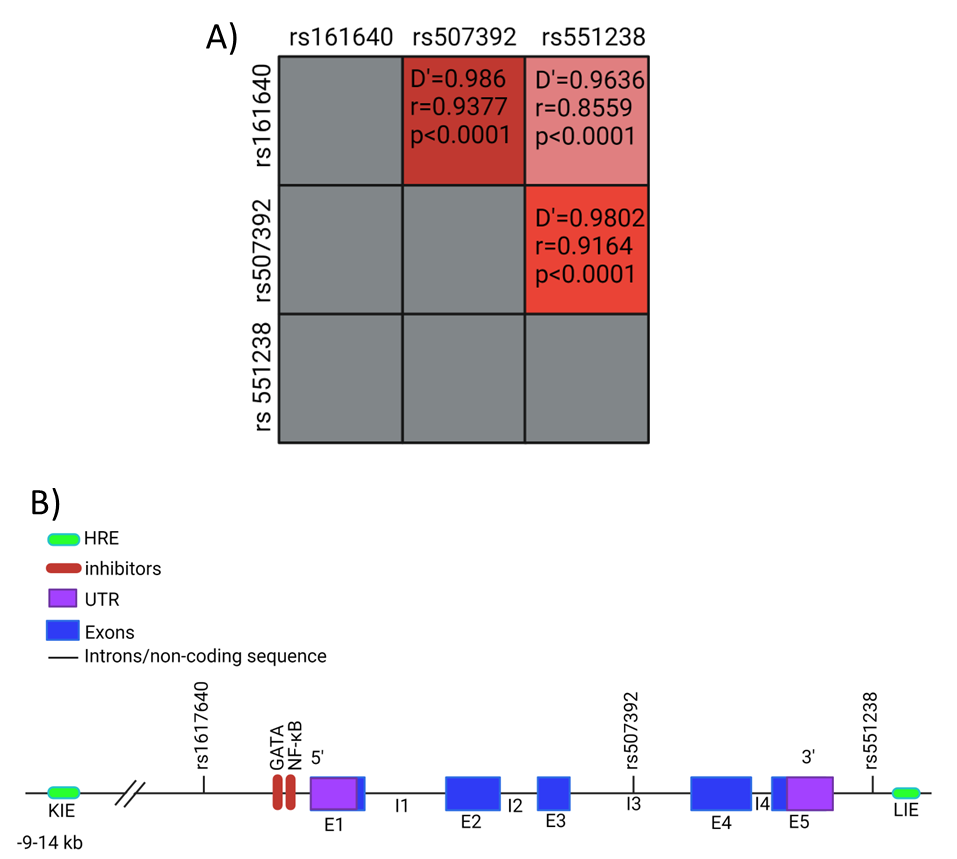

**Supplemental Figure 1 - EPO SNPs show a high degree of linkage disequilibrium.**

A) The linkage coefficient (D’), R-value and p-value are provided for each combination. Greyed boxes are duplicated combinations or analysis between the same SNPs and were therefore not analyzed. Red boxes show combinations, with darker shades indicating greater degrees of linkage disequilibrium. B) EPO gene layout with SNP locations.

**Supplemental Table 3-rs1617640 demographics and clinical parameters.**

|  | Genotype | | | p-value | | |
| --- | --- | --- | --- | --- | --- | --- |
|  | AA (n=38) | AC (n=32) | CC (n=24) | AA-AC | AA-CC | AC-CC |
| Age (years) | 64.1 ± 6.1 | 63.4 ± 6.8 | 62.6 ± 7.0 | 0.68 | 0.41 | 0.68 |
| Males (%) | 29 (76%) | 20 (63%) | 15 (63%) | 0.20 | 0.24 | 0.99 |
| Females | 9 (24%) | 12 (38%) | 9 (38%) |  |  |  |
| BMI (kg/m^2^) | 29.7 ± 5.3 | 31.8 ± 6.9 | 34.9 ± 9.3 | 0.16 | **0.007** | 0.16 |
| Normal | 7 (18%) | 5 (16%) | 2 (8%) | 0.40 | **0.008** | 0.40 |
| Pre-obese | 14 (37%) | 10 (31%) | 7 (29%) |  |  |  |
| Obese 1 | 12 (32%) | 7 (22%) | 2 (8%) |  |  |  |
| Obese 2 | 3 (8%) | 4 (13%) | 6 (25%) |  |  |  |
| Obese 3 | 2 (5%) | 6 (19%) | 7 (29%) |  |  |  |
| NYHA 1 | 8 (21%) | 6 (19%) | 4 (17%) | 0.46 | 0.67 | 0.27 |
| 2 | 17 (45%) | 9 (39%) | 12 (52%) |  |  |  |
| 3 | 8 (21%) | 11 (35%) | 6 (26%) |  |  |  |
| 4 | 5 (13%) | 5 (16%) | 1 (4%) |  |  |  |
| LVEF (%) | 58.8 ± 11.8 | 61.4 ± 10.2 | 59.2 ± 11.3 | 0.32 | 0.88 | 0.45 |
| LVEDP (mmHg) | 17.0 ± 6.7 | 17.6 ± 7.0 | 20.5 ± 5.3 | 0.72 | 0.056 | 0.13 |
| Hemoglobin (g/L) | 137.3 ± 9.1 | 135.3 ± 18.6 | 129.6 ± 15.0 | 0.56 | **0.017** | 0.24 |
| Hematocrit (L/L) | 0.40 ± 0.03 | 0.40 ± 0.05 | 0.39 ± 0.04 | 0.77 | 0.08 | 0.32 |
| MCV (fL) | 90.4 ± 3.7 | 90.2 ± 4.4 | 89.5 ± 5.0 | 0.81 | 0.44 | 0.63 |
| HbA1c (%) | 6.2 ± 1.2 | 5.9 ± 0.8 | 5.9 ± 0.7 | 0.25 | 0.31 | 0.92 |
| Cholesterol(mmol/L) | 4.2 ± 1.3 | 3.7 ± 1.1 | 4.4 ± 1.0 | 0.06 | 0.59 | **0.019** |
| HDL (mmol/L) | 1.2 ± 0.4 | 1.2 ± 0.4 | 1.3 ± 0.3 | 0.79 | 0.11 | 0.09 |
| LDL (mmol/L) | 2.2 ± 0.9 | 1.8 ± 0.9 | 2.4 ± 0.9 | 0.10 | 0.33 | **0.018** |
| LDL/HDL | 1.9 ± 0.8 | 1.6 ± 0.7 | 1.9 ± 0.8 | 0.13 | 0.80 | 0.11 |
| Non-HDL (mmol/L) | 3.0 ±1.2 | 2.5 ± 0.9 | 3.0 ± 1.0 | **0.046** | 0.99 | **0.045** |
| TG (mmol/L) | 1.9 ± 1.4 | 1.5 ± 0.7 | 1.3 ± 0.5 | 0.14 | 0.07 | 0.40 |
| RBC (x10^12/L) | 4.5 ± 0.3 | 4.4 ± 0.5 | 4.4 ± 0.4 | 0.82 | 0.32 | 0.60 |
| WBC (x10^9/L) | 7.1 ±2.1 | 7.4 ± 2.8 | 7.9 ±2.3 | 0.72 | 0.22 | 0.49 |
| Platelets (x10^9/L) | 207.0 ± 46.4 | 208.3 ± 64.6 | 199.8 ± 50.2 | 0.92 | 0.58 | 0.61 |
| Neutro (x10^9/L) | 4.6 ± 1.6 | 4.9 ± 2.7 | 5.5 ± 2.4 | 0.49 | 0.07 | 0.40 |
| Lympho (x10^9/L) | 1.7 ± 0.6 | 1.6 ± 0.6 | 1.6 ± 0.8 | 0.51 | 0.30 | 0.61 |
| NLR | 2.8 ± 1.0 | 3.5 ± 3.6 | 6.3 ± 8.7 | 0.22 | **0.018** | 0.12 |
| Troponin (ng/L) | 756.8 ± 521.6 | 796.7 ± 665.4 | 815.7 ± 599.2 | 0.79 | 0.70 | 0.91 |
| Creatine (mmol/L) | 85.2 ± 17.4 | 103.6 ± 117.8 | 92.4 ± 31.0 | 0.35 | 0.25 | 0.66 |
| Urea (mmol/L) | 6.3 ± 1.9 | 7.0 ± 2.7 | 7.1 ± 3.0 | 0.24 | 0.21 | 0.86 |

NYHA=New York Heart Association, BMI=Body mass index, HDL=high-density lipoprotein, LDL=low-density lipoprotein, LVEF=Left ventricular ejection fraction, LVEDP=left ventricular end diastolic pressure, HFpEF=Heart failure with preserved ejection fraction, T2DM=type 2 diabetes mellitus, TG=triglycerides, RBC=red blood cells, WBC=white blood cells (leukocytes), Neutro=Neutrophiles, Lympho= Lymphocytes, NLR=Neutrophiles Lymphocytes ratio. p-value for continuous variable (BMI, LVEF, LVEDP, Hemoglobin, Hematocrit, HbA1c, Cholesterol, HDL, LDL, Triglycerides, RBC, Troponin, Creatine, WBC, Neutrophiles, Lymphocytes, NLR, Urea) expressed as mean ±SD, p-value calculated using t-test. Categorical variable (Sex, BMI Class, and NYHA) expressed as number of cases (percentage of group), p-value calculated using Chi-square test. Significant p-value of <0.05 are bolded.

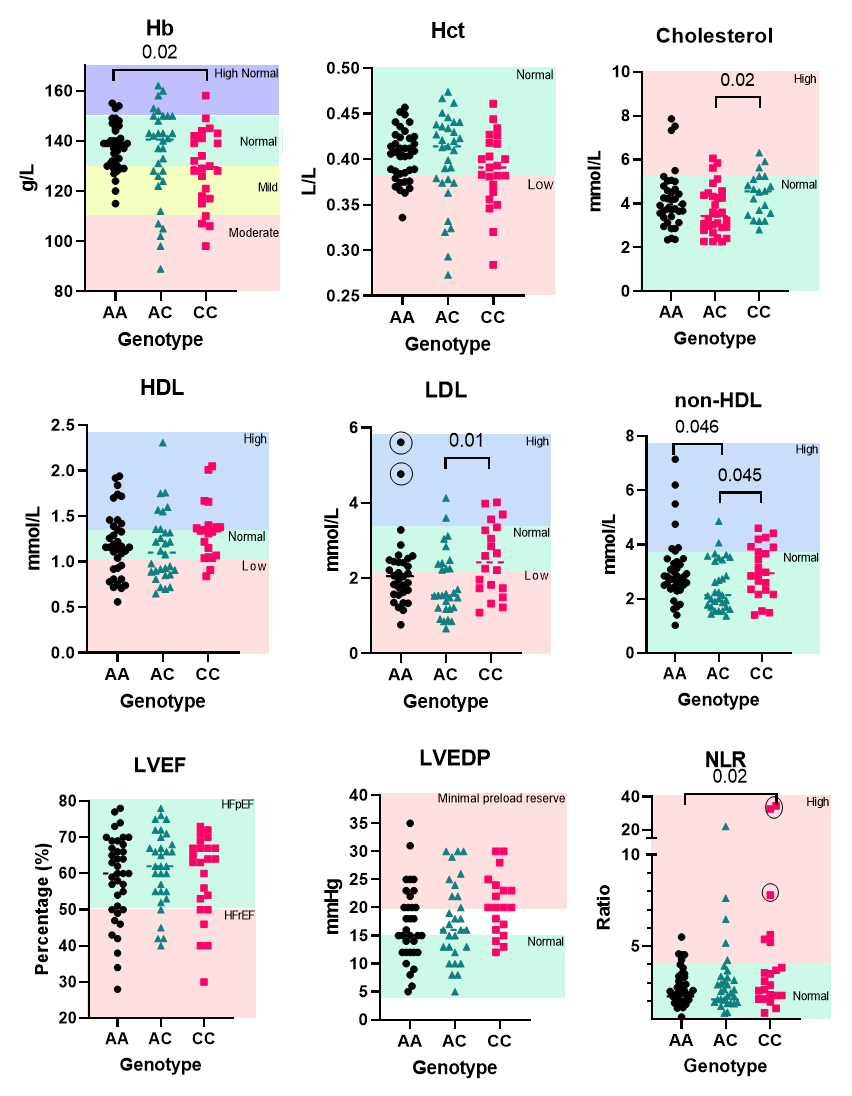

**Supplemental Figure 2- rs1617640 genotype correlation with clinical parameters.**

The clinical parameter is indicated above each graph with normal and abnormal ranges indicated within the graph by background colour. Outliers removed from statistical analysis (by ROUT test or outside limits of detection of the assay) are indicated by circles. Hb=hemoglobin, Hct=hematocrit, HDL=high-density lipoprotein, LDL=low-density lipoprotein, LVEF=left ventricular ejection fraction, LVEDP=left ventricular end diastolic pressure, and NLR=neutrophil to lymphocyte ratio. Statistical significance determined by student t-test.

**Supplemental Table 4 - rs507392 demographics and clinical parameters**

|  | Genotype | | | p-value | | |
| --- | --- | --- | --- | --- | --- | --- |
|  | AA (n=36) | AG (n=31) | GG (n=19) | AA-AG | AA-GG | AG-GG |
| Age (years) | 63.8 ± 6.1 | 63.4 ± 7.3 | 63.9 ± 6.9 | 0.89 | 0.86 | 0.80 |
| Males | 29 (81%) | 21 (68%) | 7 (37%) | 0.22 | 0.15 | 0.73 |
| Females | 7 (19%) | 10 (32%) | 12 (63%) |  |  |  |
| BMI (kg/m^2^) | 29.4 ± 5.1 | 31.1 ± 6.3 | 32.1 ± 6.6 | 0.23 | 0.10 | 0.60 |
| Normal | 7 (19%) | 5 (16%) | 2 (11%) | 0.29 | **0.036** | 0.71 |
| Pre-obese | 13 (36%) | 11 (35%) | 7 (37%) |  |  |  |
| Obese 1 | 12 (33%) | 6 (19%) | 2 (11%) |  |  |  |
| Obese 2 | 3 (8%) | 4 (13%) | 5 (26%) |  |  |  |
| Obese 3 | 1 (3%) | 5 (16%) | 3 (16%) |  |  |  |
| NYHA 1 | 8 (22%) | 6 (19%) | 4 (21%) | 0.68 | 0.79 | 0.63 |
| 2 | 15 (42%) | 10 (32%) | 9 (47%) |  |  |  |
| 3 | 8 (22%) | 11 (35%) | 5 (26%) |  |  |  |
| 4 | 5 (14%) | 4 (13%) | 1 (5%) |  |  |  |
| LVEF (%) | 59.0 ± 12.0 | 61.4 ± 10.3 | 60.5 ± 11.4 | 0.38 | 0.66 | 0.76 |
| LVEDP (mmHg) | 17.1 ± 6.7 | 17.7 ± 7.1 | 20.4 ± 4.9 | 0.75 | 0.09 | 0.18 |
| Hemoglobin (g/L) | 138.0 ± 8.8 | 137.1 ± 16.8 | 133.1 ± 13.1 | 0.78 | 0.11 | 0.39 |
| Hematocrit (L/L) | 0.41 ± 0.03 | 0.41 ± 0.05 | 0.40 ± 0.03 | 0.96 | 0.32 | 0.51 |
| MCV (fL) | 90.7 ± 3.4 | 90.4 ± 4.0 | 90.5 ± 4.0 | 0.75 | 0.82 | 0.96 |
| HbA1c (%) | 6.2 ± 1.2 | 5.9 ± 0.8 | 5.9 ± 0.7 | 0.20 | 0.25 | 0.92 |
| Cholesterol (mmol/L) | 4.2 ± 1.3 | 3.7 ± 1.0 | 4.6 ± 0.9 | 0.07 | 0.33 | **0.005** |
| HDL (mmol/L) | 1.2 ± 0.4 | 1.2 ± 0.4 | 1.3 ± 0.3 | 0.86 | 0.14 | 0.11 |
| LDL (mmol/L) | 2.2 ± 0.9 | 1.8 ± 0.8 | 2.6 ± 0.9 | 0.11 | 0.16 | **0.005** |
| LDL/HDL | 1.9 ± 0.8 | 1.6 ± 0.7 | 2.0 ± 0.7 | 0.15 | 0.51 | 0.051 |
| Non-HDL (mmol/L) | 3.1 ± 1.2 | 2.5 ± 0.9 | 3.2 ± 0.9 | **0.046** | 0.63 | **0.011** |
| TG (mmol/L) | 1.9 ± 1.4 | 1.4 ± 0.7 | 1.4 ± 0.6 | 0.057 | 0.11 | 0.98 |
| RBC (x10^12/L) | 4.5 ± 0.3 | 4.5 ± 0.5 | 4.4 ± 0.4 | 0.92 | 0.54 | 0.63 |
| WBC (x10^9/L) | 7.2 ± 2.1 | 7.1 ± 2.4 | 7.9 ±2.3 | 0.88 | 0.25 | 0.25 |
| Platelets (x10^9/L) | 208.0 ± 47.5 | 206.2 ± 63.2 | 202.4 ± 52.1 | 0.89 | 0.69 | 0.83 |
| Neutrophiles (x10^9/L) | 4.6 ± 1.7 | 4.7 ± 2.3 | 5.5 ± 2.4 | 0.85 | 0.09 | 0.22 |
| Lymphocytes (x10^9/L) | 1.8 ± 0.6 | 1.7 ± 0.6 | 1.6 ± 0.8 | 0.56 | 0.48 | 0.78 |
| NLR | 2.8 ± 1.0 | 3.3 ± 3.6 | 6.3 ± 9.4 | 0.36 | **0.031** | 0.12 |
| Troponin (ng/L) | 782.2 ± 523.4 | 795.3 ± 663.0 | 859.6 ± 631.2 | 0.93 | 0.64 | 0.74 |
| Creatine (mmol/L) | 86.1 ± 17.4 | 81.8 ± 25.2 | 91.6 ± 30.1 | 0.42 | 0.40 | 0.23 |
| Urea (mmol/L) | 6.4 ± 1.9 | 6.4 ± 1.7 | 7.2 ± 2.9 | 0.99 | 0.23 | 0.23 |

NYHA=New York Heart Association, BMI=Body mass index, HDL=high-density lipoprotein, LDL=low-density lipoprotein, LVEF Left ventricular ejection fraction, LVEDP=left ventricular end diastolic pressure, HFpEF=Heart failure with preserved ejection fraction, T2DM=type 2 diabetes mellitus, TG=triglycerides, RBC=red blood cells, WBC=white blood cells (leukocytes), Neutro=Neutrophiles, Lympho= Lymphocytes, NLR=Neutrophiles Lymphocytes ratio. p-value for continuous variable (BMI, LVEF, LVEDP, Hemoglobin, Hematocrit, HbA1c, Cholesterol, HDL, LDL, Triglycerides, RBC, Troponin, Creatine, WBC, Neutrophiles, Lymphocytes, NLR, Urea) expressed as mean ±SD, p-value calculated using student t-test. Categorical variable (Sex, BMI Class, and NYHA expressed as number of cases (percentage of group), p-value calculated using Chi-square test. Significant p-value of <0.05 are bolded.

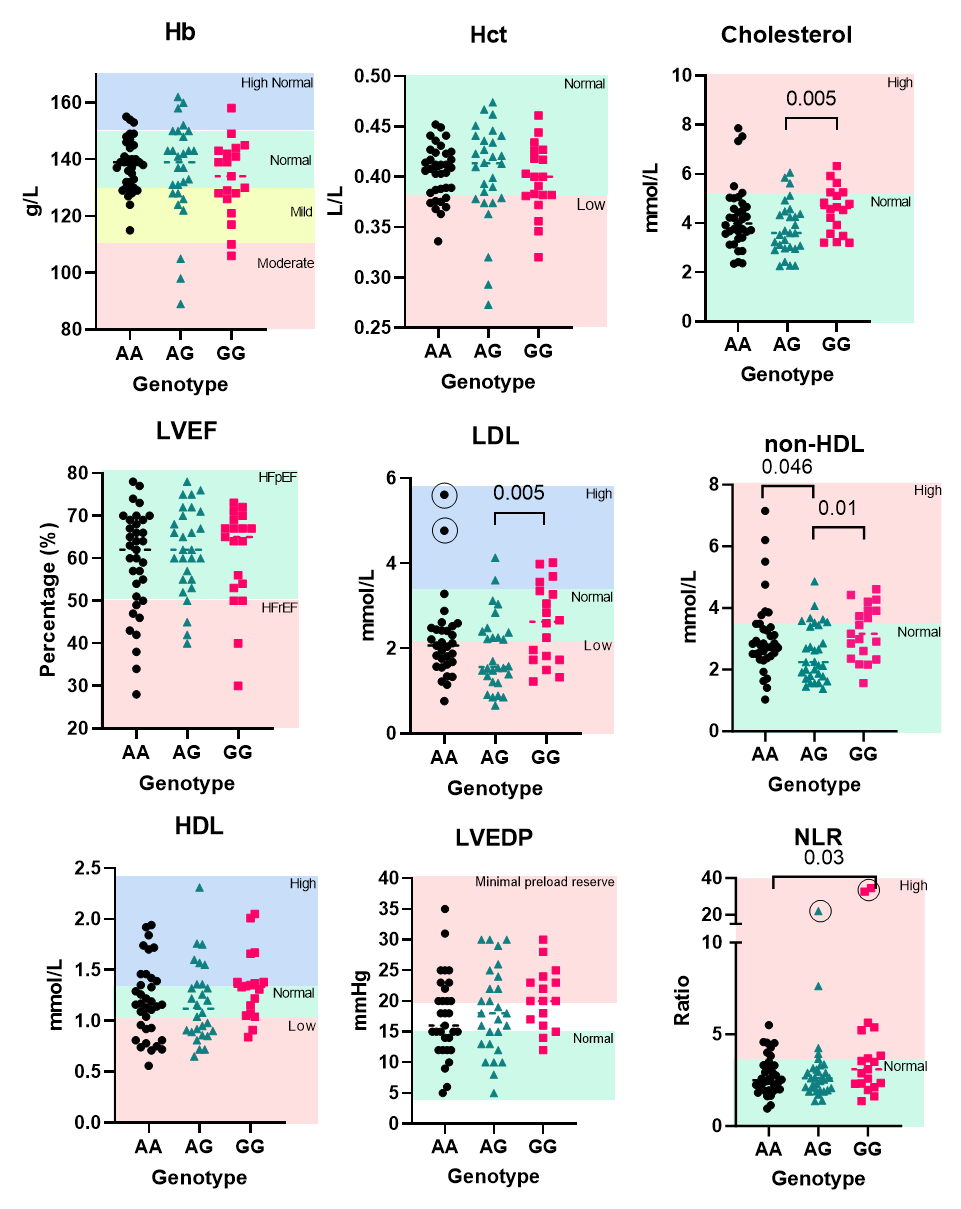

**Supplemental Figure 3- rs507392 genotype correlations with clinical parameters.**

Clinical parameters are indicated above each graph, with normal and abnormal ranges indicated within the graph by background colour. Outliers removed from statistical analysis (by ROUT test or outside limits of detection of the assay) are indicated by circles. Hb=hemoglobin, Hct=hematocrit, HDL=high-density lipoprotein, LDL=low-density lipoprotein, LVEF=left ventricular ejection fraction, LVEDP=left ventricular end diastolic pressure, and NLR=neutrophile to lymphocyte ratio. Statistical significance was determined by student t-test.

**Supplemental Table 5 - rs551238 demographics and clinical parameters**

|  | Genotypes | | | p-value | | |
| --- | --- | --- | --- | --- | --- | --- |
|  | TT | TG | GG | TT-TG | TT-GG | TG-GG |
| Age (years) | 63.9 ± 6.0 | 63.4 ± 7.1 | 63.6 ± 7.3 | 0.74 | 0.89 | 0.90 |
| Males | 32 (80%) | 20 (65%) | 11 (65%) | 0.14 | 0.21 | 0.98 |
| Females | 8 (20%) | 11 (35%) | 6 (35%) |  |  |  |
| BMI (kg/m^2^) | 30.0 ± 6.0 | 31.3 ± 6.6 | 32.9 ± 6.5 | 0.38 | 0.11 | 0.44 |
| Normal | 7 (18%) | 5 (16%) | 2 (12%) | 0.19 | 0.06 | 0.19 |
| Pre-obese | 14 (35%) | 11 (35%) | 5 (29%) |  |  |  |
| Obese 1 | 14 (35%) | 5 (16%) | 2 (12%) |  |  |  |
| Obese 2 | 3 (8%) | 4 (13%) | 5 (29%) |  |  |  |
| Obese 3 | 2 (5%) | 6 (19%) | 3 (18%) |  |  |  |
| NYHA 1 | 9 (23%) | 7 (23%) | 2 (12%) | 0.88 | 0.83 | 0.83 |
| 2 | 16 (41%) | 10 (32%) | 8 (47%) |  |  |  |
| 3 | 9 (23%) | 9 (29%) | 6 (35%) |  |  |  |
| 4 | 5 (13%) | 5 (16%) | 1 (6%) |  |  |  |
| LVEF (%) | 59.7 ± 11.8 | 61.7 ± 10.1 | 58.9 ± 11.5 | 0.45 | 0.82 | 0.39 |
| LVEDP (mmHg) | 16.8 ± 6.5 | 18.1 ± 7.1 | 19.9 ± 5.1 | 0.42 | 0.11 | 0.42 |
| Hemoglobin (g/L) | 139.1 ± 9.4 | 136.0 ± 15.5 | 131.9 ± 15.6 | 0.31 | 0.04 | 0.40 |
| Hematocrit (L/L) | 0.41 ± 0.03 | 0.40 ± 0.04 | 0.39 ± 0.04 | 0.48 | 0.11 | 0.46 |
| MCV (fL) | 90.8 ± 3.3 | 89.8 ± 4.3 | 91.0 ± 4.1 | 0.29 | 0.83 | 0.36 |
| HbA1c (%) | 6.3 ± 1.3 | 5.9 ± 0.8 | 5.9 ± 0.8 | 0.17 | 0.26 | 0.97 |
| Cholesterol (mmol/L) | 4.2 ± 1.2 | 3.7 ± 1.1 | 4.4 ± 1.0 | 0.06 | 0.57 | **0.031** |
| HDL (mmol/L) | 1.2 ± 0.4 | 1.2 ± 0.4 | 1.3 ± 0.3 | 0.77 | 0.13 | 0.09 |
| LDL (mmol/L) | 2.2 ± 0.9 | 1.9 ± 0.9 | 2.4 ± 0.9 | 0.14 | 0.45 | 0.06 |
| LDL/HDL | 1.9 ± 0.8 | 1.6 ± 0.6 | 1.9 ± 0.8 | 0.12 | 0.97 | 0.21 |
| Non-HDL (mmol/L) | 3.1 ± 1.2 | 2.5 ± 0.9 | 3.1 ± 1.0 | **0.045** | 0.97 | 0.06 |
| TG(mmol/L) | 1.9 ± 1.4 | 1.4 ± 0.7 | 1.4 ± 0.5 | 0.07 | 0.18 | 0.88 |
| RBC (x10^12/L) | 4.5 ± 0.3 | 4.5 ± 0.5 | 4.4 ± 0.5 | 0.83 | 0.16 | 0.35 |
| WBC (x10^9/L) | 7.2 ± 2.1 | 7.1 ± 2.4 | 8.0 ± 2.5 | 0.89 | 0.23 | 0.25 |
| Platelets (x10^9/L) | 206.6 ± 46.0 | 211.9 ± 63.5 | 188.6 ± 47.4 | 0.68 | 0.19 | 0.20 |
| Neutrophiles (x10^9/L) | 4.6 ± 1.6 | 4.7 ±2.3 | 5.6 ± 2.6 | 0.76 | 0.08 | 0.23 |
| Lymphocytes (x10^9/L) | 1.8 ± 0.6 | 1.6 ± 0.6 | 1.6 ± 0.8 | 0.32 | 0.35 | 0.83 |
| NLR | 2.7 ± 1.0 | 3.5 ± 3.6 | 6.7 ± 9.9 | 0.22 | **0.018** | 0.11 |
| Troponin (ng/L) | 769.9 ± 505.5 | 858.2 ± 704.0 | 822.6 ± 563.5 | 0.56 | 0.73 | 0.86 |
| Creatine (mmol/L) | 85.5 ± 17.0 | 80.8 ± 24.8 | 94.6 ± 31.3 | 0.35 | 0.17 | 0.10 |
| Urea (mmol/L) | 6.4 ± 1.9 | 6.3 ± 1.9 | 7.5 ± 3.0 | 0.81 | 0.12 | 0.12 |

NYHA=New York Heart Association, BMI=Body mass index, HDL=high-density lipoprotein, LDL=low-density lipoprotein, LVEF=Left ventricular ejection fraction, LVEDP=left ventricular end diastolic pressure, HFpEF=Heart failure with preserved ejection fraction, T2DM=type 2 diabetes mellitus, TG=triglycerides, RBC=red blood cells, WBC=white blood cells (leukocytes), Neutro=Neutrophiles, Lympho= Lymphocytes, NLR=Neutrophiles Lymphocytes ratio. p-value for continuous variable (BMI, LVEF, LVEDP, Hemoglobin, Hematocrit, HbA1c, Cholesterol, HDL, LDL, Triglycerides, RBC, Troponin, Creatine, WBC, Neutrophiles, Lymphocytes, NLR, Urea) expressed as mean ±SD, p-value calculated using t-test. Categorical variable (Sex, BMI Class, and NYHA) expressed as number of cases (percentage of group), p-value calculated using Chi-square test. Significant p-value of <0.05 are bolded.

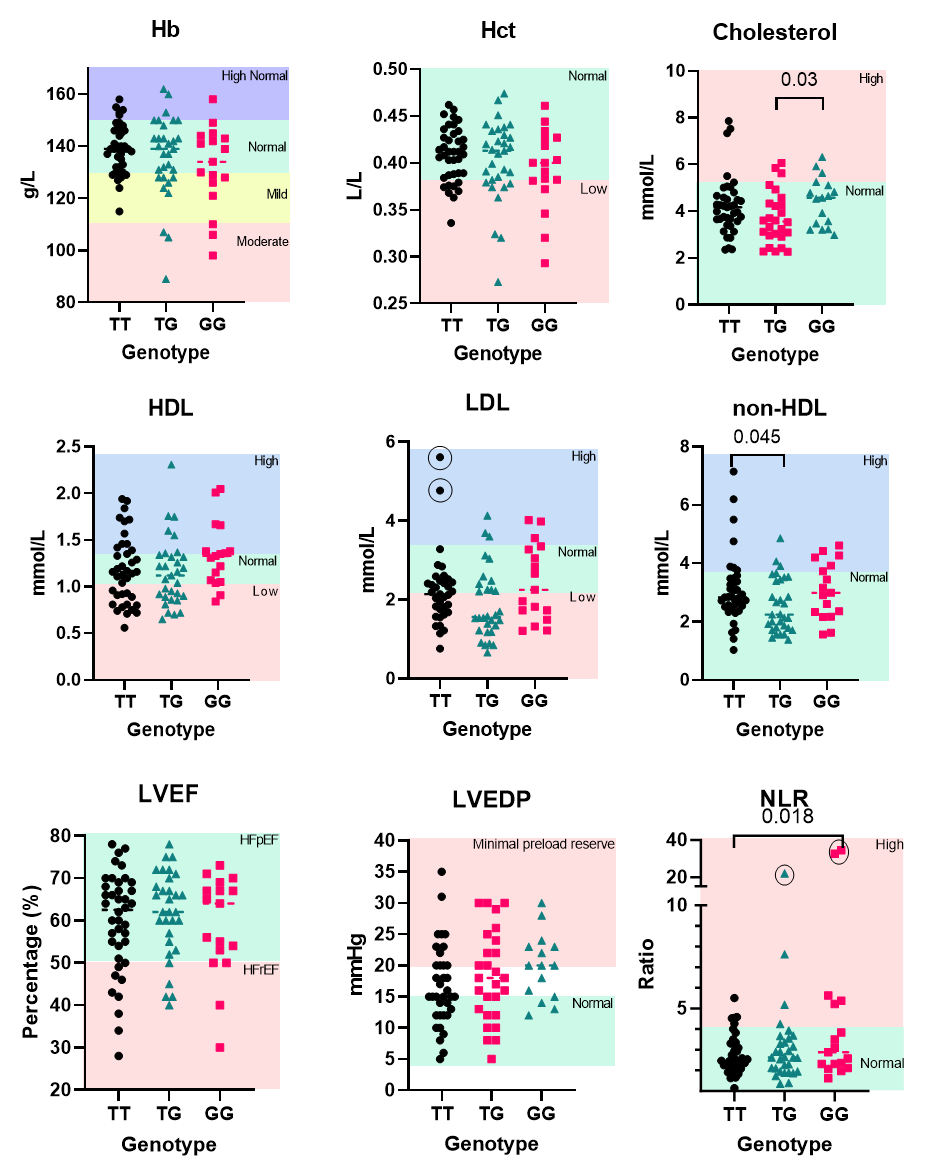

**Supplemental Figure 4 - rs551238 genotype correlations with clinical parameters.**

The clinical parameter is indicated above each graph with normal and abnormal ranges indicated within the graph by background colour. Outliers removed from statistical analysis (by ROUT test or outside limits of detection) are indicated by circles. Hb=hemoglobin, Hct=hematocrit, HDL=high-density lipoprotein, LDL=low-density lipoprotein, LVEF=left ventricular ejection fraction, LVEDP=left ventricular end diastolic pressure, and NLR=neutrophile to lymphocyte ratio. Statistical significance determined by student t-test.

**Supplemental Table 6- Haplotype associations with clinical phenotypes.**

| Haplotype | HFpEF | | | T2DM | | | |
| --- | --- | --- | --- | --- | --- | --- | --- |
|  | Frequency | OR (95% CI) | p-value | | Frequency | OR (95% CI) | p-value |
| AAT | 0.5674 | 1.00 | n/a | 0.5673 | | 1.00 | n/a |
| CGG | 0.3826 | 1.51 (0.64-3.56) | 0.35 | 0.3844 | | 0.99 (0.51-1.91) | 0.97 |
| CGT | 0.292 | 0.68 (0.02-27.15) | 0.84 | 0.0301 | | 0.26 (0.02-2.82) | 0.27 |
| GHA p-value | 0.34 | | | 0.8 | | | |

GHA=global haplotype association, T2DM=Type 2 Diabetes mellitus, HFpEF=Heart Failure with preserved ejection fraction, OR=Odds ratio

**Supplemental Table 7- Sex differences in SNPs correlations with phenotypes**

|  | | T2DM | | | | HFpEF | | | |
| --- | --- | --- | --- | --- | --- | --- | --- | --- | --- |
| S | Geno. | Cases | Controls | OR (95% CI) | Int. | Cases | Controls | OR (95% CI) | Int. |
| rs1617640 | | | | | | | | | |
| F | A/A | 1 | 8 | 1.00 | 0.8 | 7 | 2 | 1.00 | 0.48 |
|  | A/C | 4 | 8 | 4.83 (0.37-63.55) |  | 9 | 3 | 1.01 (0.11-9.48) |  |
|  | C/C | 1 | 8 | 0.80 (0.03-18.12) |  | 8 | 1 | 2.54 (0.15-43.18) |  |
| M | A/A | 10 | 19 | 1.00 |  | 23 | 6 | 1.00 |  |
|  | A/C | 11 | 9 | 1.96 (0.56-6.88) |  | 18 | 2 | 2.94 (0.48-18.14) |  |
|  | C/C | 5 | 10 | 0.61 (0.14-2.64) |  | 11 | 4 | 1.13 (0.13-4.48) |  |
| rs507392 | | | | | | | | | |
| F | A/A | 1 | 6 | 1.00 | 0.97 | 5 | 2 | 1.00 | 0.48 |
|  | A/G | 2 | 8 | 1.13 (0.06-21.74) |  | 9 | 1 | 3.62 (0.20-66.26) |  |
|  | G/G | 1 | 6 | 0.45 (0.01-14.55) |  | 7 | 0 | n/a |  |
| M | A/G | 10 | 19 | 1.00 |  | 23 | 6 | 1.00 |  |
|  | G/G | 11 | 10 | 1.61 (0.46-5.67) |  | 18 | 3 | 1.90 (0.37-9.73) |  |
|  | C/C | 4 | 8 | 0.63 (0.13-3.02) |  | 10 | 2 | 2.01 (0.27-15.06) |  |
| rs551238 | | | | | | | | | |
| F | T/T | 1 | 7 | 1.00 | 0.9 | 5 | 3 | 1.00 | 0.26 |
|  | T/G | 3 | 8 | 2.79 (0.18-43.94) |  | 10 | 1 | 7.30 (0.48-111.05) |  |
|  | G/G | 1 | 5 | 1.14 (0.04-32.91) |  | 6 | 0 | n/a |  |
| M | T/T | 12 | 20 | 1.00 |  | 26 | 6 | 1.00 |  |
|  | T/G | 10 | 10 | 1.40 (0.40-4.92) |  | 17 | 3 | 1.67 (0.32-8.67) |  |
|  | G/G | 4 | 7 | 0.63 (0.13-3.10) |  | 9 | 2 | 1.78 (0.23-13.69) |  |

S=sex, F=female, M=male, Geno.=genotype, OR=odds ratio, Ctrl.=controls, int=interaction p-value, T2DM=Type 2 diabetes mellites, HFpEF=Heart failure with preserved ejection fraction

**Supplemental Table 8 - Other variants identified by Sanger sequencing**

| Variants | Total | Anemia | Dyslipidemia | T2DM | HFpEF | Hypertension |
| --- | --- | --- | --- | --- | --- | --- |
| -681C>T | 1 | 0 | 0 | 0 | 1 | 0 |
| -591C>T | 12 | 4 | 8 | 7 | 11 | 9 |
| -500G>T | 1 | 1 | 1 | 1 | 0 | 1 |
| -494T>G | 1 | 1 | 1 | 1 | 0 | 1 |
| -454T>C | 1 | 1 | 0 | 0 | 0 | 0 |
| -433T>A | 1 | 1 | 0 | 0 | 0 | 0 |
| -428G>A | 1 | 1 | 0 | 0 | 1 | 1 |
| -422G>A | 2 | 1 | 1 | 0 | 1 | 1 |
| 673A>G | 1 | 1 | 1 | 1 | 1 | 1 |
| 1775A>G | 2 | 0 | 2 | 2 | 1 | 2 |
| 1789insA | 3 | 1 | 2 | 2 | 3 | 3 |
| 1797insA | 1 | 0 | 0 | 0 | 0 | 1 |
| 1817G>A | 4 | 1 | 4 | 3 | 4 | 4 |
| 1826G>A | 1 | 0 | 1 | 1 | 1 | 1 |
| 2159C>T | 6 | 3 | 3 | 1 | 6 | 4 |
| 2163C>T | 6 | 3 | 3 | 1 | 6 | 4 |
| 2167?-2170?del^1^ | 76 | 19 | 56 | 24 | 63 | 57 |
| 2167?-2170?del^2^ | 4 | 1 | 3 | 3 | 4 | 4 |
| 2237G>A | 1 | 0 | 1 | 1 | 1 | 1 |
| 2374C>A | 1 | 0 | 1 | 0 | 0 | 0 |
| 2376insC | 1 | 1 | 1 | 0 | 1 | 1 |
| 2384C>A | 1 | 0 | 1 | 0 | 1 | 1 |
| 2397C>T | 1 | 0 | 1 | 1 | 0 | 1 |
| 2404G>A | 1 | 1 | 1 | 1 | 1 | 1 |
| 3107C>T | 3 | 1 | 2 | 1 | 3 | 2 |
| 3132insG | 1 | 1 | 1 | 0 | 1 | 1 |
| 3136G>A | 1 | 0 | 1 | 0 | 0 | 1 |
| 3149C>G | 1 | 0 | 0 | 0 | 1 | 1 |
| 3150T>A | 1 | 0 | 1 | 0 | 0 | 1 |
| 3154T>G | 77 | 20 | 55 | 26 | 62 | 56 |
| 3154T>G | 6 | 1 | 3 | 4 | 6 | 5 |
| 3194G>C | 1 | 0 | 1 | 1 | 1 | 1 |
| 3289T>A | 1 | 0 | 1 | 1 | 1 | 1 |
| 3300T>A | 1 | 0 | 1 | 1 | 1 | 1 |
| 3324G>A | 1 | 0 | 1 | 1 | 1 | 1 |
| 3363insT | 1 | 0 | 1 | 0 | 0 | 1 |
| 3434C>T | 4 | 1 | 3 | 0 | 3 | 3 |
| 3573C>A | 1 | 0 | 1 | 1 | 1 | 1 |
| 3606insG | 1 | 0 | 1 | 1 | 1 | 1 |
| 3786C>T | 1 | 0 | 1 | 0 | 0 | 1 |
| 3793insC | 1 | 0 | 1 | 0 | 0 | 1 |
| 3798C>T | 1 | 1 | 0 | 0 | 1 | 0 |
| 3806T>C | 2 | 0 | 2 | 0 | 2 | 2 |
| 3806T>C | 31 | 9 | 23 | 12 | 26 | 25 |
| 3833insA | 1 | 0 | 1 | 0 | 0 | 1 |
| 3747insA | 2 | 0 | 1 | 1 | 2 | 2 |
| 3892G>A | 2 | 0 | 1 | 0 | 1 | 1 |
| 3892insC | 1 | 0 | 1 | 0 | 0 | 1 |
| 3948T>G | 1 | 0 | 1 | 0 | 1 | 1 |
| 3976G>T | 1 | 1 | 0 | 1 | 1 | 1 |

ins=insertion, del=deletion, T2DM=type 2 diabetes mellites, HFpEF=heart failure with preserved ejection fraction

^1-^deletion is heterozygous

^2-^deletion is homozygous
